## Supplementary material for "Refractory Inflammatory Arthritis definition and model generated through patient and multi-disciplinary professional modified Delphi process": S1 - Key Qual Findings

**Supplementary Figure S1:** Key components of RD and Persistent Symptoms identified by Patients and Rheumatologists from initial thematic analysis

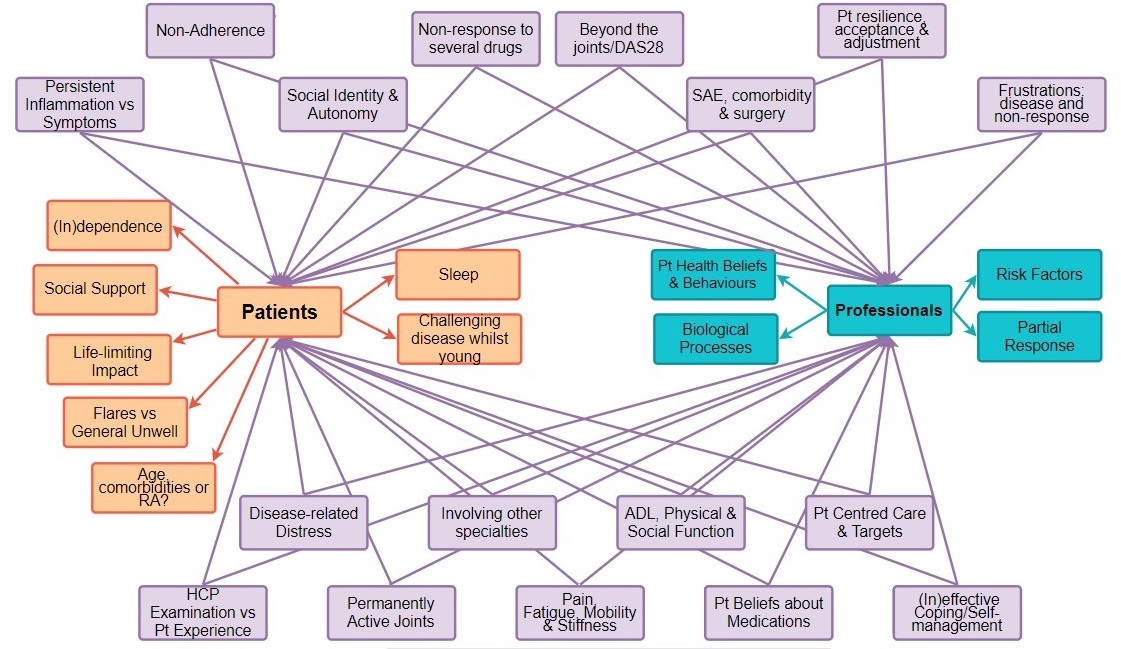

Please note: Patient only components in orange, Healthcare Professional only components in blue, with shared components in purple.

Patient interviews were conducted with 20 people living with RA (including one partner) and five adults with PolyJIA interviewed from four UK Rheumatology Outpatient departments, who were either currently on bDMARD not responding with DAS28>3.2 (n=21), or DAS28>2.6 if PGA≥50 (n=4). Patients had previous experience with on average 3 csDMARDs and 3.5 bDMARDs, with mean MSK-HQ=24.6, mean DAS28=4.60, and mean Patient Global=55.4. Healthcare professional interviews were conducted with 32 HCPs from 11 UK Hospitals interviewed (average 11.7 years of Rheumatology experience, range 1-29), representing Rheumatology (Consultant, Registrar), Clinical Specialist Nursing, Psychology, Physiotherapy, Occupational Therapy, Podiatry, Pharmacy and Social Work.
