## Supplementary material for "Refractory Inflammatory Arthritis definition and model generated through patient and multi-disciplinary professional modified Delphi process": S2 - Components lists

**Supplementary Data S2:** Lists of Components and Domains presented during a) Nominal Group, b) Round One Online Voting, and c) Round Two Online Voting

**S2a:** Initial list of Components (n=72) and Domains (n=12) presented during to Nominal Group

1. Pain
   1.1 Pain during the day
   1.2 Pain during the night
   1.3 Pain when active
   1.4 Pain when resting
2. Stiffness
   2.1 Stiffness during the day (lasting longer than 30 minutes in the morning)
   2.2 Stiffness during the night
   2.3 Stiffness when active
   2.4 Stiffness when resting
3 Fatigue/low energy
  3.1 Lack of physical energy making it difficult to conduct daily activities e.g. washing, dressing
  3.2 Lack of mental energy making it difficult to concentrate and remember things
  3.3 Feeling fatigued after being active for short or long periods
  3.4 Fatigue lasting several hours or all day
4 Poor Quality sleep
  4.1 Difficulty falling asleep
  4.2 Difficulty staying asleep
  4.3 Sleeping too much
  4.4 Sleeping too little
  4.5 Waking unrefreshed
5 Overall Disease Activity
  5.1 Overall high rating of disease activity/impact
  5.2 Unpredictable disease course
  5.3 Variable disease course – relapsing, remitting
6 Joint Activity/Involvement
  6.1 Joint replacement(s) or fusion(s)
  6.2 Accrued damage - Joint erosion(s), deformity(ies) or restrictions in range of movement (may or may not be painful)
  6.3 One or two persistently active/affected joints despite good control in other joints
  6.4 Disease activity in previously unaffected joints e.g. new joint swelling
  6.5 Involvement of joints outside of DAS28 e.g. feet, ankles, hips and TMJ
7 Co-morbidities and Wider Involvement outside of Joints (previous or new)
  7.1 Presence of Co-mobidity(ies) e.g. Cardiovascular Disease
  7.2 Extra-articular Manifestation(s) e.g. Interstitial Lung Disease
  7.3 Involvement of other inflammatory features outside of DAS28 e.g. Uveitis or Vasculitis
  7.4 Involvement of other non-inflammatory features outside of DAS28 e.g. muscle weakness
  7.5 Feeling systemically/generally unwell
  7.6 Problematic side-effects e.g. from medications or other illnesses/infections
8 Steroid Use and Dependency
  8.1 Repeated need of short course steroid tablets or intra-articular injections, that may or may not control flare and localised swelling
  8.2 Inability to taper steroid tablets longer term (steroid dependency)
9 Experiences of DMARD Inefficacy, Intolerability or Discontinuation (single or multiple occurrences)
  9.1 Primary inefficacy – no response to DMARD at all
  9.2 Secondary inefficacy – developed resistance to DMARD over time
  9.3 Poor tolerability of DMARD due to side effects particularly infections or administration
  9.4 Temporary or Permanent discontinuation of DMARD due to illness or surgery
  9.5 Experience of combinations of inefficacy, intolerability or discontinuation
10 Physical Functioning and Quality of Life
   10.1 Inability to perform desired activities e.g. hobbies, social, school, work
   10.2 Reduced mobility e.g. driving, use of public transport, needing to sit most of the day
   10.3 Problems walking, standing or climbing stairs
   10.4 Trouble bending, lifting or reaching things from above
   10.5 Problems with self-care e.g. washing/dressing
   10.6 Needing help from others for activities of daily living e.g. partner/family or paid carer for household management, getting dressed or running errands
   10.7 Poor physical function – strength, dexterity, grip
   10.8 Unable to rely on previous body functions/abilities
   10.9 Current Physical Activity Level not at desired level
11 Psychological Functioning and Quality of Life
   11.1 Disease-related distress e.g. psychological distress related to burden of disease
        11.1.1 Physical related Distress – distress from pain, fatigue, loss of function or disability
        11.1.2 Emotional Distress – Acceptance, Disease Burden, Unpredictability, worry about prognosis or impact on relationships
        11.1.3 Social Distress – Impact on personal relationships
        11.1.4 Treatment Distress - Fear of surgery, Fear/Anxiety of trying new treatments and/or side-effects
        11.1.5 Healthcare Distress – frustrations with healthcare professionals, services and processes
   11.2 Depression
        11.2.1 Low mood
        11.2.2 Loss of interest/pleasure in activities e.g. hobbies, socialising, activities of daily living
   11.3 Anxiety
        11.3.1 Worrying about different things or far future problems e.g pregnancy, salaried work
        11.3.2 Unable to relax easily
        11.3.3 Feeling tense or nervous
   11.4 Low Self-Efficacy/Esteem/Confidence – Perceived ability to manage and cope with symptoms
   11.5 Cognitive Impairments e.g. brain fog or orientation or perceptual functions
   11.6 Poor understanding of Illness or low health literacy
12 Social Functioning and Quality of Life
   12.1 Reduced independence or autonomy
   12.2 Increased dependence on others to engage socially e.g. reliance on partner to attend events
   12.3 Social activity limitations e.g. difficulties in executing social activities
   12.4 Restriction of social participation e.g. involvement in life situations
   12.5 Reduced Social Support Network or Relationship breakdowns
   12.6 Isolation
   12.7 Loneliness
   12.8 Barriers of physical, social and attitudinal environment in which people live and conduct their lives – negative attitudes of others, no adjustments made to housing, inflexible working

**S2b:** Full final list of Components (n=73) and Domains (n=10) presented during Round One Online Voting

1 Pain
  1.1 Pain during the day
  1.2 Pain during the night
  1.3 Pain when active
  1.4 Pain when resting
  1.5 Pain in joints e.g. hands and feet
  1.6 Pain in other areas e.g. muscles, neuropathic, regional (e.g. back), widespread etc
2 Joint Stiffness
  2.1 Joint stiffness during the day (lasting longer than 30 minutes in the morning)
  2.2 Joint stiffness during the night
  2.3 Joint stiffness when active
  2.4 Joint stiffness when resting
3 Fatigue/low energy
  3.1 Lack of physical energy resulting in difficulties conducting daily activities e.g. washing, dressing
  3.2 Lack of mental energy leading to difficulties with concentration and memory
  3.3 Feeling fatigued following physical activities over short or long periods
  3.4 Feeling fatigued irrespective of physical activities e.g. fatigue upon waking
  3.5 Fatigue lasting several hours or all day
4 Poor sleep
  4.1 Difficulty falling asleep
  4.2 Difficulty staying asleep
  4.3 Sleeping too much (>10 hours/day)
  4.4 Sleeping too little (<5 hours/day)
  4.5 Poor quality sleep
  4.6 Waking up unrefreshed
5 Overall Disease Activity
  5.1 Overall high rating of disease activity/impact from high inflammatory markers at a single appointment
  5.2 Overall high rating of disease activity/impact from high patient global score at a single appointment
  5.3 Persistently high inflammation and/or symptoms e.g. for at least two consecutive clinical visits over the period of at least six months
  5.4 Persistent high inflammation and/or symptoms with only minimal fluctuations e.g. for at least two consecutive clinical visits over the period of at least six months
  5.5 Persistent high inflammation and/or symptoms with large fluctuations e.g. for at least two consecutive clinical visits over the period of at least six months
  5.6 Unpredictable or variable disease course – relapsing-remitting with periods of both high and low activity
6 Joint Activity/Involvement
  6.1 Joint replacement(s) or fusion(s) due to inflammation
  6.2 Accrued damage due to inflammation - Joint erosion(s), deformity(ies) or restrictions in range of movement (may or may not be painful)
  6.3 Accrued damage, joint erosion(s), replacement(s) or fusion(s) due to Osteoarthritis
  6.4 One or two persistently active/affected joints despite acceptable control in other joints
  6.5 Disease activity in previously unaffected joints e.g. new joint swelling or pain
  6.6 Feet and/or ankle(s) involvement
  6.7 Involvement of joint activity outside of DAS28 e.g. hips, TMJ, tendonitis or enthesitis
7 Co-morbidities and Wider Involvement outside of Joints (previous or new)
  7.1 Presence of Extra-articular Manifestation(s), Complications or associated pathology of Inflammatory Arthritis e.g. Secondary Sjögrens, Nodules, Interstitial Lung Disease, (JIA-associated) Uveitis, Cardiovascular Disease, Anaemia or Chronic leg ulcers
  7.2 Presence of OA and FM as secondary Co-morbidity(ies)
  7.3 Presence of other Co-morbidity(ies) e.g. non-Hodgkin’s lymphoma and osteoporosis, Asthma, Diabetes, high blood pressure or Depression
  7.4 Involvement of other inflammatory features outside of DAS28 e.g. Uveitis or Vasculitis
  7.5 Involvement of other non-inflammatory features outside of DAS28 e.g. muscle weakness or cachexia
  7.6 Feeling systemically/generally unwell e.g. fever, sweats
  7.7 Problematic side-effects e.g. from medications or other illnesses/infections
8 Healthcare and Medication Utilisation related to Inflammatory Arthritis/Symptoms
  8.1 GP appointment(s)
  8.2 Hospital Accident and Emergency visit, admission(s) or inpatient stay
  8.3 Hospital outpatient appointment(s)
  8.4 Additional use of rheumatology service outside of scheduled appointments e.g. extra calls to the helpline or contact with the rheumatology nurse
  8.5 New prescriptions
  8.6 Multiple prescribed medications, including high dose analgesics
  8.7 Use of Complementary and Alternative Medicine
  8.8 Repeated need of short course steroid tablets or intra-articular injections, that may or may not control flare and localised swelling
  8.9 Inability to taper steroid tablets longer term (steroid dependency)
  8.10 Dependence on analgesics, particularly opioids
9 Experiences of DMARD Inefficacy, Intolerability or Discontinuation
  9.1 Primary inefficacy – no response to DMARD at all
  9.2 Secondary inefficacy – developed ‘resistance’ to DMARD over time
  9.3 Poor tolerability of DMARD due to side effects particularly infections, nausea or administration
  9.4 Temporary or Permanent discontinuation of DMARD due to illness, infection, surgery or toxicity
  9.5 Experience of multiple occurrences of inefficacy, intolerability or discontinuation
10 Functioning and Quality of Life – Physical, Psychological and Social
   10.1 Inability to perform desired activities e.g. hobbies, social, salaried/voluntary work
   10.2 Reduced mobility e.g. driving, use of public transport, needing to sit most of the day
   10.3 Problems walking, standing or climbing stairs
   10.4 Problems with self-care e.g. washing/dressing
   10.5 Poor physical function e.g. lack of strength, dexterity, grip
   10.6 Problems with Sexual Functioning and Relationships
   10.7 Pain Interference impacting on quality of life
   10.8 Disease-related distress e.g. psychological distress related to burden of disease including Physical related, Emotional, Social, Treatment or Healthcare Distress
   10.9 General Emotional Distress e.g. suspected or diagnosed Depression or Anxiety
   10.10 Low Self-Efficacy/Esteem/Confidence e.g. perceived low ability to manage and cope with symptoms
   10.11 Cognitive Impairments e.g. brain fog, orientation or perceptual functions
   10.12 Poor understanding of Illness or low health literacy
   10.13 Reduced independence/autonomy or Increased dependence on others for activities of daily living
   10.14 Reduced Social Support Network, Relationship breakdowns/difficulties or restriction of social participation
   10.15 Isolation e.g. quantity of relationships or from unemployment, or Loneliness e.g. perception of disagreement between actual and desired levels of social contact or perceived quality of relationships
   10.16 Barriers in physical, social and attitudinal environment e.g. negative attitudes of other people without the condition such as discrimination and lack of understanding/empathy, no adjustments made to housing, inflexible working
   10.17 Negative effect on education, work (paid/voluntary), or accessing welfare benefits

**S2c:** Full list of Components (n=13) presented during Round Two Online Voting

1 Co-morbidities and Wider Involvement Outside of Joints (Comorbid)
  1.1 Presence of Extra-articular Manifestation(s), Complications or associated pathology of Inflammatory Arthritis e.g. Secondary Sjögrens, Nodules, Interstitial Lung Disease, (JIA-associated) Uveitis, Cardiovascular Disease, Anaemia or Chronic leg ulcers
  1.2 Involvement of other inflammatory features outside of DAS28 e.g. Uveitis or Vasculitis Poor tolerability of DMARD due to side effects particularly infections, nausea or administration
  1.3 Involvement of other non-inflammatory features outside of DAS28 e.g. muscle weakness or cachexia
  1.4 Disease Activity not captured by DAS28 (hands, shoulders, wrists, elbows and knees) including involvement of other joints (hips, TMJ, feet), other inflammatory features (vasculitis, uveitis, tendonitis or enthesitis) or non-inflammatory features (muscle weakness or cachexia)
2 Poor Sleep (Sleep)
  2.1 Waking up unrefreshed
  2.2 Poor Quality Sleep due to Inflammatory Arthritis
3 Healthcare and Medications Utilisation (HCMed)
  3.1 Inability to taper steroid tablets longer term (steroid dependency)
  3.2 Repeated need of short course steroid tablets or intra-articular injections, that may or may not control flare and localised swelling
  3.3 Dependence on analgesics, particularly opioids
4 Exclusion criteria - Flare Indicators (Exclusion)
  4.1 Overall high rating of disease activity/impact from high inflammatory markers at a single appointment
  4.2 Overall high rating of disease activity/impact from high patient global score at a single appointment
  4.3 Disease activity in previously unaffected joints e.g. new joint swelling or pain
  4.4 Hospital Accident and Emergency visit, admission(s) or inpatient stay due to Inflammatory Arthritis
