## Supplementary figures and images for "Refractory Inflammatory Arthritis definition and model generated through patient and multi-disciplinary professional modified Delphi process"

### S3 - Example of Voting Questions

**Supplementary Figure S3:** Example of Delphi Voting Questions (Round One Online)

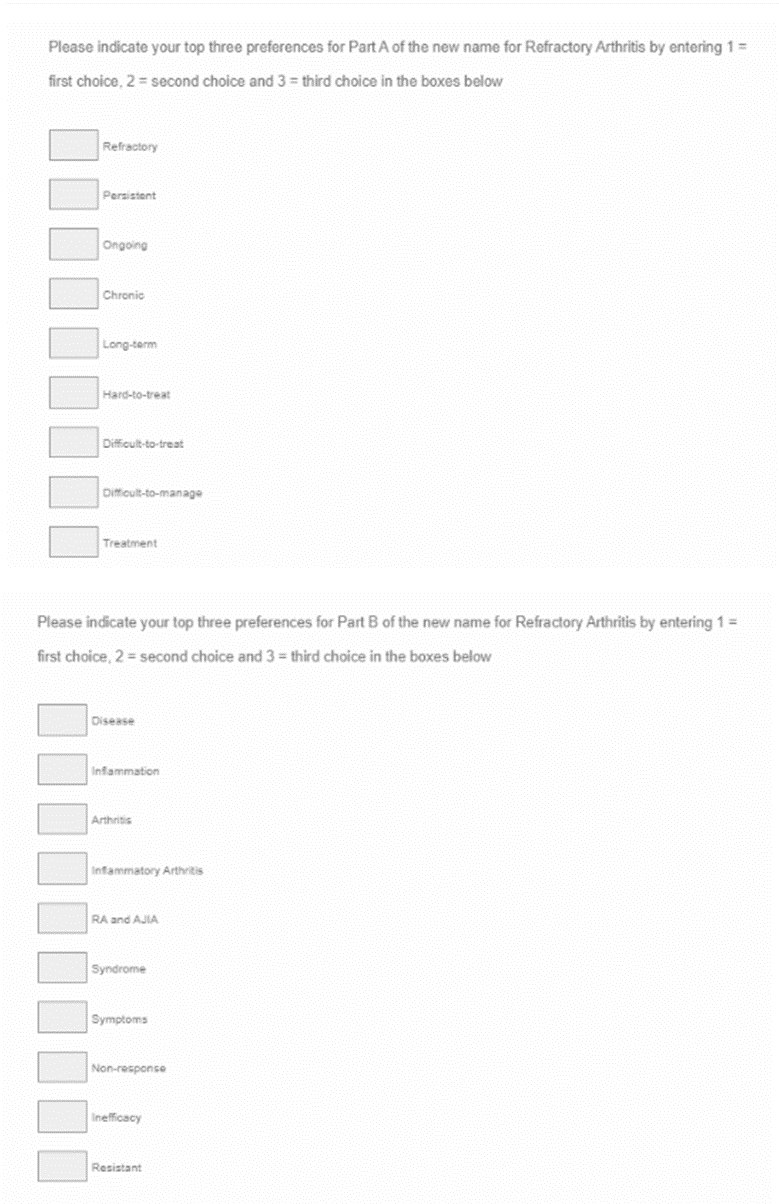

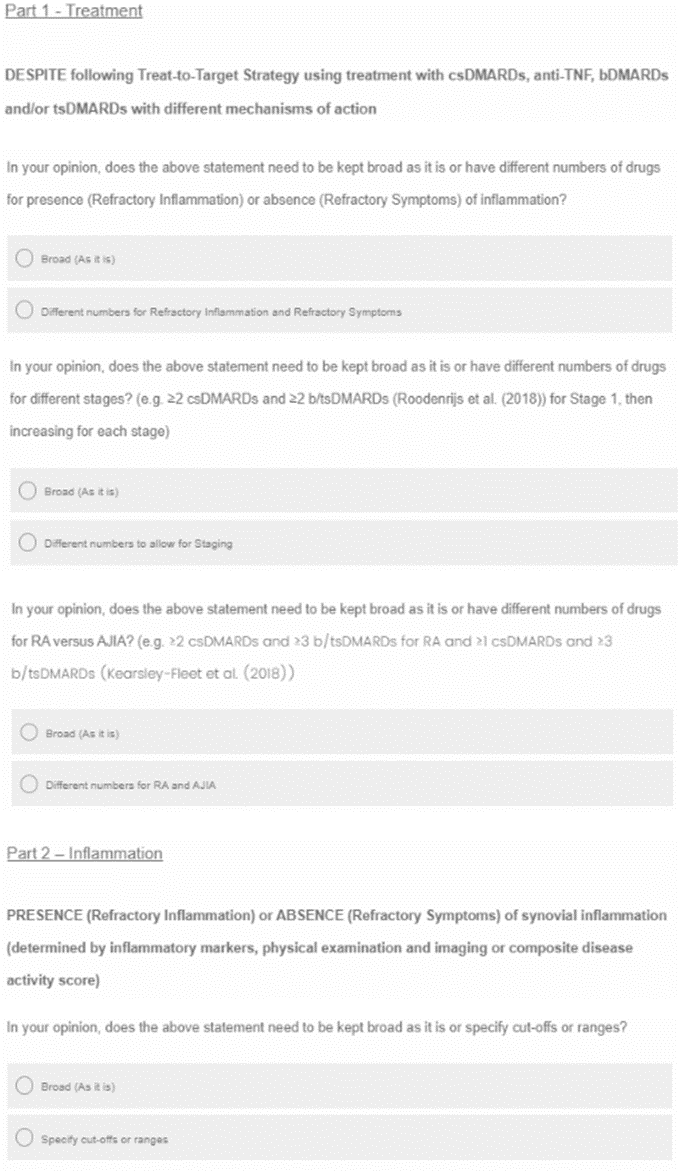

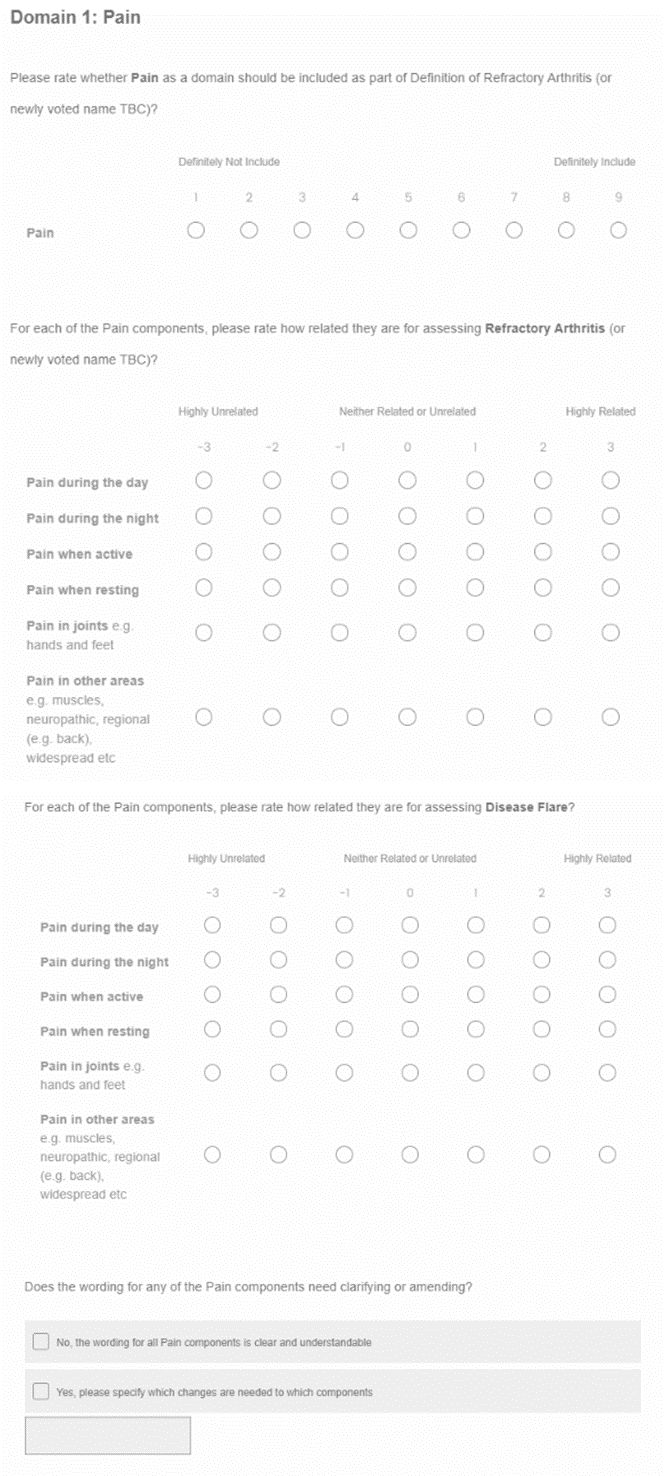

### S6 - Definition Development Overview

**Supplementary Figure S6:** Overview of Definition Development and Refinement

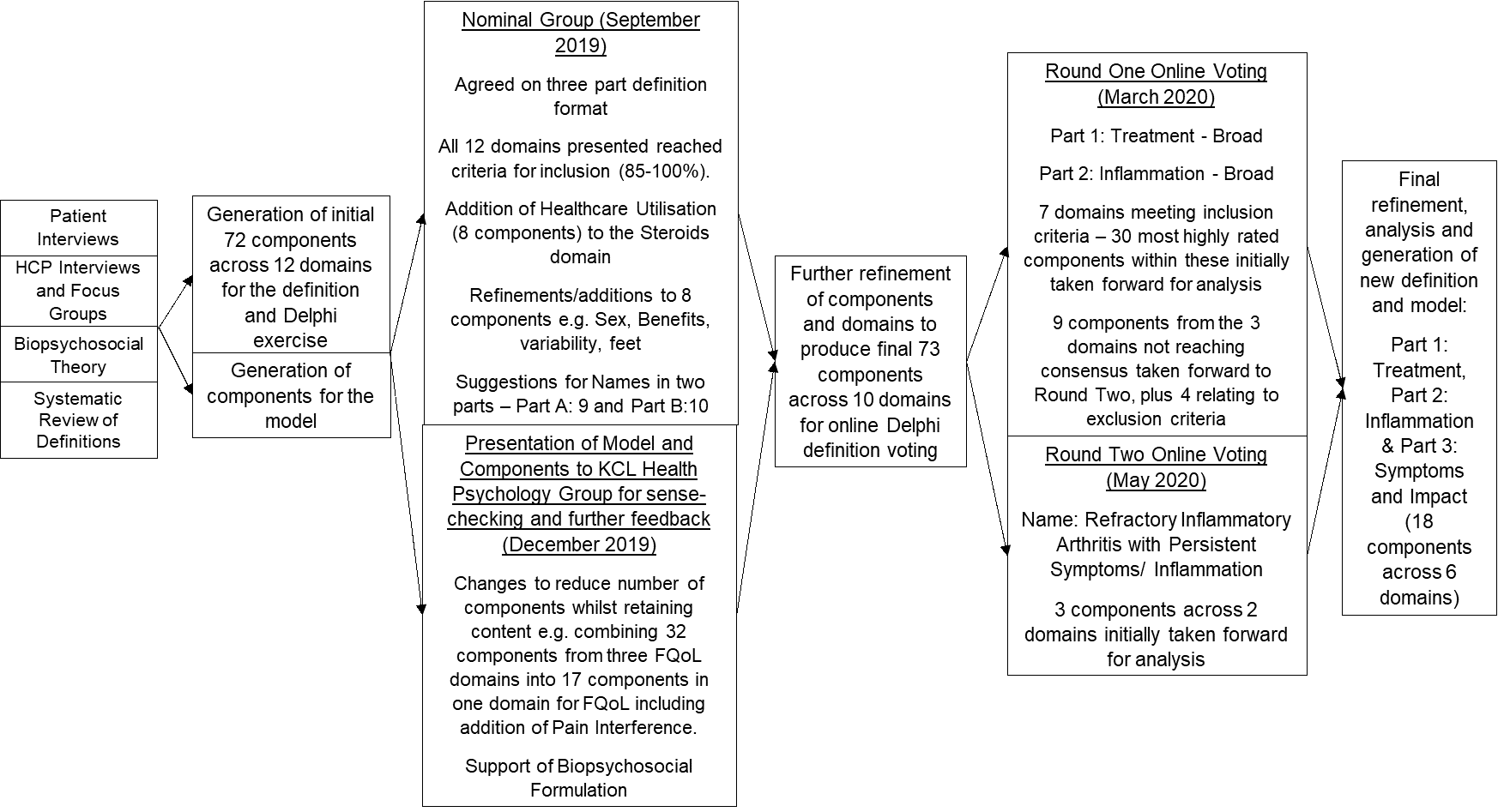
