## Supplementary material for "Refractory Inflammatory Arthritis definition and model generated through patient and multi-disciplinary professional modified Delphi process": S4 - Participants across rounds

**Supplementary Data S4:** Analysis of Participants across Delphi Rounds

There were no differences in participant characteristics (e.g. Rheumatology experience or diversity of roles) between each round for those that completed, nor between those who dropped out (n=3 each online round). There was no difference in years of Rheumatology experience between participants who started each Delphi Round as determined by one-way ANOVAs either in total (F(2,103) = 0.14, *p* = .871) or within each Role Group (Rheumatologists: F(2,30) = 0.52, *p* = .603, Patients: F(2,18) = 0.29, *p* = .752, Other HCPs: F(2,39) = 0.45, *p* = .641 and Researchers: F(2,7) = 1.01, *p* = .411). There was no difference in the diversity of different roles involved in each Delphi Round as determined by Fishers exact tests, for either proportions represented at the broader Role Group level (*p* = .378) nor specific individual Roles (*p* = .996). Finally there was no difference in the training/working areas of participants as determined by Fishers exact tests, as those experienced in Adult, Paediatric or Both were no different across rounds (*p* = .664) nor levels of participants who had received specific MSK training or not (*p* = .571).

There was no difference in years of Rheumatology experience between participants who fully completed each Delphi Round as determined by one-way ANOVAs either in total (F(2,97) = 0.06, *p* = .944) or within each Role Group (Rheumatologists: F(2,28) = 0.20, *p* = .817, Patients: F(2,17) = 0.15, *p* = .859, Other HCPs: F(2,36) = 0.33, *p* = .721 and Researchers: F(2,7) = 0=1.01, *p* = .411). There was no difference in the diversity of different roles who fully completed each Delphi Round as determined by Fishers exact tests, for either proportions represented at the broader Role Group level (*p* = .392) nor specific individual Roles (*p* = .995). Finally there was no difference in the training/working areas of participants as determined by Fishers exact tests, as those experienced in Adult, Paediatric or Both were no different across rounds in those for fully completed voting (*p* = .544) nor levels of participants who had received specific MSK training or not (*p* = .595).
