## Supplementary material for "Refractory Inflammatory Arthritis definition and model generated through patient and multi-disciplinary professional modified Delphi process": S5 - Name Preference Voting

**Supplementary Table S5:** Options for Name preference voting with Proportions and 95% Confidence Intervals

| Nominal Group (n=12)† | | Online Round 1 (n=40) | | | | Online Round 2 (n=53) | |
| --- | --- | --- | --- | --- | --- | --- | --- |
| Name | Proportion of Votes [95%CIs] | Part A Name | Proportion of Votes [95%CIs] | Part B Name | Proportion of Votes [95%CIs] | Name | Proportion of Votes [95%CIs] |
| Refractory disease | 0.42 [0.20, 0.63] | A1 Refractory | 0.26 [0.20, 0.32] | B1 Disease | 0.25 [0.19, 0.30] | Refractory Disease | 0.14 [0.10, 0.17] |
| Persistent disease | 0.21 [0.03, 0.39] | A2 Persistent | 0.23 [0.17, 0.28] | B2 Inflammation | 0.07 [0.04, 0.11] | Refractory Arthritis | 0.11 [0.08, 0.15] |
| Difficult to treat | 0.04 [-0.06, 0.14] | A3 Ongoing | 0.04 [0.01, 0.07] | B3 Arthritis | 0.22 [0.17, 0.34] | Refractory Inflammatory Arthritis | 0.25 [0.20, 0.29] |
| Chronic disease | 0.04 [-0.06, 0.14] | A4 Chronic | 0.06 [0.03, 0.09] | B4 Inflammatory Arthritis | 0.19 [0.14, 0.24] | Persistent Disease | 0.08 [0.05, 0.11] |
| Treatment non-responsive | 0.04 [-0.06, 0.14] | A5 Long-term | 0.02 [0.00, 0.03] | B5 RA and AJIA | 0.13 [0.08, 0.17] | Persistent Arthritis | 0.08 [0.05, 0.11] |
| Treatment Inefficacy | 0.04 [-0.06, 0.14] | A6 Hard-to-treat | 0.08 [0.05, 0.12] | B6 Syndrome | 0.02 [0.00, 0.03] | Persistent Inflammatory Arthritis | 0.19 [0.15, 0.24] |
| Treatment failure | 0.00 [-0.02, 0.02] | A7 Difficult-to-treat | 0.17 [0.12, 0.22] | B7 Symptoms | 0.09 [0.05, 0.13] | Difficult-to-treat Disease | 0.03 [0.01, 0.06] |
| Treatment resistant | 0.13 [-0.03, 0.28] | A8 Difficult-to-manage | 0.12 [0.07, 0.16] | B8 Non-response | 0.01 [-0.01, 0.02] | Difficult-to-treat Arthritis | 0.04 [0.02, 0.06] |
| Hard to treat | 0.00 [-0.02, 0.02] | A9 Treatment | 0.03 [0.00, 0.05] | B9 Inefficacy | 0.00 [0.00, 0.00] | Difficult-to-treat Inflammatory Arthritis | 0.08 [0.04, 0.11] |
| Difficult to manage | 0.08 [-0.05, 0.21] |  |  | B10 Resistant | 0.03 [0.01, 0.06] |  |  |

†Note: one missing voter due to late arrival
