## Supplementary material for "Refractory Inflammatory Arthritis definition and model generated through patient and multi-disciplinary professional modified Delphi process": S7 - Statistics for Components

**Supplementary Tables S7:** A) Statistics for Round One Components Considered for Definition

| Domain and Components Considered | Statistics (As stated and [95%CIs]) | Levels of Agreement | Justification | Final Decision |
| --- | --- | --- | --- | --- |
| 1 - Disease Activity | Percentage Include: 94.8% [82.7%, 99.3%]  Mean Include: 8.28 [7.96, 8.61] | Level: Strong |  |  |
| Persistently high inflammation and/or symptoms e.g. for at least two consecutive clinical visits over the period of at least six months | Mean Related: 2.56 [2.37, 2.76]  Mean diff: 0.92, *d*: 0.67 | I-CVI: 0.95 (Excellent) | Large mean diff with medium-to-large effect size. Highest scoring Ref score overall (total and across groups) (R1 – Disease Activity 3) | Definition  Combined as:  Persistently high inflammation and/or symptoms e.g. with or without fluctuations for at least two consecutive clinical visits over the period of at least six months |
| Persistent high inflammation and/or symptoms with only minimal fluctuations e.g. for at least two consecutive clinical visits over the period of at least six months | Mean Related: 2.41 [2.15, 2.68]  Mean diff: 0.90, *d*: 0.62 | I-CVI: 0.90 (Excellent) | Large mean diff with medium-to-large effect size. High scoring Ref score (total and across groups) (R1 – Disease Activity 4) |  |
| Persistent high inflammation and/or symptoms with large fluctuations e.g. for at least two consecutive clinical visits over the period of at least six months | Mean Related: 1.97 [1.63, 2.32]  Mean diff: 0.54, *d*: 0.38 | I-CVI: 0.69 (Good) | Small mean diff with medium effect size. High scoring Ref score (total and across groups) but lowest of the three(R1 – Disease Activity 5) |  |
| 2 - Joint Stiffness | Percentage Include: 92.3% [79.1%, 98.4%]  Mean Include: 8.21 [7.83, 8.58] | Level: Strong |  |  |
| Joint stiffness during the day (lasting longer than 30-60 minutes in the morning) | Mean Related: 2.54 [2.31, 2.77]  Mean diff: 0.18, *d:* 0.18 | I-CVI: 0.92 (Excellent) | High related score for this domain (total and for all groups) but low mean diff with Flare. Highest rated component for Rheum (R1 – Stiffness 1). Free text Comment: consider extending to 60 minutes | Definition |
| Joint stiffness when resting | Mean Related: 1.69 [1.34, 2.04]  Mean diff: 0.05, *d*: 0.04 | I-CVI: 0.60 (Good) | Medium related score (total and for all groups) but low mean diff with Flare (R1 – Stiffness 4). Free text comments: Several had issues with the wording | Model |

| Domain and Components Considered | Statistics (As stated and [95%CIs]) | Levels of Agreement | Justification | Final Decision |
| --- | --- | --- | --- | --- |
| 3 - Joint Activity | Percentage Include: 89.47% [75.2%, 97.1%] Mean Include: 8.21 [7.83, 8.59] | Level: Strong |  |  |
| One or two persistently active/affected joints despite acceptable control in other joints | Mean Related: 2.00 [1.69, 2.31]  Mean diff: 1.11, *d*: 0.78 | I-CVI: 0.68 (Good) | Large mean diff with large effect size. Highest scoring Ref related score for the domain (totals, Rheums and patients) (R1 – Joint Activity 4) | Definition |
| Involvement of joint activity outside of DAS28 e.g. hips, TMJ, tendonitis or enthesitis | Mean Related: 1.82 [1.44, 2.19]  Mean diff: 0.16, *d*: 0.12 | I-CVI: 0.65 (Good) | High scoring Ref related score (totals, Rheums and patients) but very small mean diff with Flare (R1 – Joint Activity 7) | Included in R2 voting as Comorbid 4 |
| Accrued damage due to inflammation - Joint erosion(s), deformity(ies) or restrictions in range of movement (may or may not be painful) | Mean Related: 1.37 [0.85, 1.88]  Mean diff: 1.68, *d:* 0.95 | I-CVI: 0.54 (Fair) | Largest mean diff score with large effect size but low-to-medium related Ref score (total). Voted highly related by patients (R1 – Joint Activity 2) | Definition |
| Joint replacement(s) and/or fusion(s) due to inflammation | Mean Related: 0.79 [0.21, 1.37]  Mean diff: 1.26, *d*: 0.68 | I-CVI: 0.37 (Poor) | Large mean diff with medium-to-large effect size but very low related Ref score (total). Medium rated by patients and HCPs (R1 – Joint Activity 1) | Model |
| 4 - Pain | Percentage Include: 82.1% [66.5%, 92.5%]  Mean Include: 7.79 [7.27, 8.32] | Level: Strong |  |  |
| Pain in joints e.g. hands and feet | Mean Related: 2.38 [2.13, 2.64]  Mean diff: -0.08, *d*: -0.10 | I-CVI: 0.87 (Excellent) | Highest related score for this domain (total and for all groups) but very low negative mean diff with Flare. Highest rated for Rheum for this domain (R1 – Pain 5) | Definition |

| Domain and Components Considered | Statistics (As stated and [95%CIs]) | Levels of Agreement | Additional Comments | Final Decision |
| --- | --- | --- | --- | --- |
| 4 – Pain (Continued) |  |  |  |  |
| Pain during the day | Mean Related: 2.10 [1.80, 2.41]  Mean diff: 0.03, *d*: 0.03 | I-CVI: 0.85 (Excellent) | High related score (total and for all groups) but very low mean diff with Flare (R1 – Pain 1) | Definition  Combined as:  Pain during the day and/or night |
| Pain in other areas e.g. muscles, neuropathic, regional (e.g. back), widespread etc | Mean Related: 0.85 [0.28, 1.42]  Mean diff: 0.36, *d:* 0.20 | I-CVI: 0.39 (Poor) | Very highly rated for Patients but not Rheum (unrelated) – Biggest diff with Flare (R1 – Pain 6) | Model |
| 5 – Fatigue | Percentage Include: 79.5% [63.5%, 90.7%]  Mean Include: 7.54 [6.92, 8.16] | Level: Moderate |  |  |
| Lack of physical energy resulting in difficulties conducting daily activities e.g. washing, dressing | Mean Related: 2.05 [1.70, 2.40]  Mean diff: 0.18, *d*: 0.16 | I-CVI: 0.74 (Excellent) | Highest related score for this domain (total and for all groups) (R1 – Fatigue 1) | Definition |
| Fatigue lasting several hours or all day | Mean Related: 1.85 [1.42, 2.27]  Mean diff: 0.05, *d*: 0.04 | I-CVI: 0.63 (Good) | High related score (total and for patients and HCPs) but very low mean diff with Flare (R1 – Fatigue 5) | Model |
| Lack of mental energy leading to difficulties with concentration and memory | Mean Related: 1.82 [1.45, 2.19]  Mean diff: 0.28, *d*: 0.20 | I-CVI: 0.63 (Good) | High related score (total and for patients and HCPs) but biggest mean diff with Flare (R1 – Fatigue 2) | Definition |
| 6 - Functioning and Quality of Life | Percentage Include: 78.4% [61.8%, 90.2%]  Mean Include: 7.78 [7.30, 8.26] | Level: Moderate |  |  |
| 6a - Physical |  |  |  |  |
| Pain Interference impacting on quality of life | Mean Related: 2.16 [1.81, 2.52]  Mean diff: 0.27, *d*: 0.22 | I-CVI: 0.76 (Excellent) | Highest related component for this domain (totals and all groups) (R1 – FQoL PPS 7) | Definition  Moved to Pain Domain |
| Problems with self-care e.g. washing/dressing | Mean Related: 2.11 [1.71, 2.51]  Mean diff: 0.05, *d*: 0.04 | I-CVI: 0.73 (Good) | Highest overall rated component for Patients and highly rated by others (R1 – FQoL PPS 4) | Definition |

| Domain and Components Considered | Statistics (As stated and [95%CIs]) | Levels of Agreement | Justification | Final Decision |
| --- | --- | --- | --- | --- |
| 6 - Functioning and Quality of Life (Continued) |  |  |  |  |
| 6a – Physical (Continued) |  |  |  |  |
| Inability to perform desired activities e.g. hobbies, social, salaried/voluntary work | Mean Related: 2.03 [1.62, 2.43]  Mean diff: 0.00, *d:* 0.00 | I-CVI: 0.67 (Good) | High related score (total and for all groups) but same score as Flare. Highly related for other HCPs (R1 – FQoL PPS 1) | Definition |
| Poor physical function e.g. lack of strength, dexterity, grip | Mean Related: 2.03 [1.64, 2.42]  Mean diff: 0.05, *d*: 0.05 | I-CVI: 0.73 (Good) | High related score (total and for all groups) but very small mean diff with Flare. Highly related for Patients (R1 – FQoL PPS 5) | Definition |
| Reduced mobility e.g. driving, use of public transport, needing to sit most of the day | Mean Related: 2.00 [1.62, 2.38]  Mean diff: 0.00, *d*: 0.00 | I-CVI: 0.73 (Good) | High related score (total and for all groups) but same score as Flare (R1 – FQoL PPS 2) | Definition  Combined as:  Reduced mobility and/or Problems walking, standing, or climbing stairs e.g. driving, use of public transport, needing to sit |
| Problems walking, standing or climbing stairs | Mean Related: 2.00 [1.62, 2.38]  Mean diff: 0.00, *d*: 0.00 | I-CVI: 0.73 (Good) | High related score (total and for all groups) but same score as Flare (R1 – FQoL PPS 3) |  |
| 6b – Psychological |  |  |  |  |
| Disease-related distress e.g. psychological distress related to burden of disease including Physical, Emotional, Social, Treatment/Healthcare Distress | Mean Related: 2.08 [1.72, 2.44]  Mean diff: 0.41, *d*: 0.32 | I-CVI: 0.70 (Good) | Highest related component for this domain but low mean diff with flare. Rated equally highly by Patients and other HCPs (R1 – FQoL PPS 8) | Definition |
| General Emotional Distress e.g. suspected or diagnosed Depression or Anxiety | Mean Related: 1.65 [1.18, 2.12]  Mean diff: 0.65, *d:* 0.44 | I-CVI: 0.52 (Fair) | Medium mean diff with medium effect size and medium Ref related score (R1 – FQoL PPS 9) | Model |
| Low Self-Efficacy/Esteem/Confidence e.g. perceived low ability to manage and cope with symptoms | Mean Related: 1.27 [0.79, 1.75]  Mean diff: 0.54, *d:* 0.36 | I-CVI: 0.35 (Poor) | Small mean diff with small-to-medium effect size and medium Ref related score (R1 – FQoL PPS 10) | Model |
| 6c – Social |  |  |  |  |
| Reduced Social Support Network, Relationship breakdowns/difficulties or restriction of social participation | Mean Related: 1.11 [0.55, 1.66]  Mean diff: 0.76, *d:* 0.43 | I-CVI: 0.36 (Poor) | Medium mean diff with medium effect size and medium Ref related score (R1 – FQoL PPS 14) | Model |

| Domain and Components Considered | Statistics (As stated and [95%CIs]) | Levels of Agreement | Justification | Final Decision |
| --- | --- | --- | --- | --- |
| 6 - Functioning and Quality of Life (Continued) |  |  |  |  |
| 6c – Social (Continued) |  |  |  |  |
| Isolation e.g. quantity of relationships or from unemployment, or Loneliness e.g. perception of disagreement between actual and desired levels of social contact or perceived quality of relationships | Mean Related: 1.03 [0.44, 1.61]  Mean diff: 0.73, *d:* 0.40 | I-CVI: 0.37 (Poor) | Medium mean diff with medium effect size and small/medium Ref related score (R1 – FQoL PPS 15) | Model |
| 7 – DMARD Experiences | Percentage Include: 73% [55.9%, 86.2%]  Mean Include: 7.41 [6.70,8.11] | Level: Moderate |  |  |
| Primary inefficacy (no response to DMARD at all) | Mean Related: 2.27 [1.83, 2.71]  Mean diff: 1.49, *d*: 0.85 | I-CVI: 0.81 (Excellent) | Large mean diff with large effect size and large Ref related score (R1 – DMARD 1) | Definition  Combined as:  Primary inefficacy (no response to DMARD at all) and/or Secondary inefficacy (developed DMARD resistance over time) |
| Secondary inefficacy (developed ‘resistance’ to DMARD over time) | Mean Related: 2.14 [1.70, 2.57]  Mean diff: 0.73, *d*: 0.50 | I-CVI: 0.78 (Excellent) | Medium mean diff with medium effect size and large Ref related score (R1 – DMARD 2) |  |
| Experience of multiple occurrences of inefficacy, intolerability or discontinuation | Mean Related: 2.00 [1.47, 2.53]  Mean diff: 1.46, *d*: 0.75 | I-CVI: 0.76 (Excellent) | Large mean diff with large effect size and large Ref related score. Agreement between Rheum and Patients (R1 – DMARD 5) | Definition |

Please note that domains and some components were rated on whether to be included (1 = ‘Definitely Not Include’ to 9 = ‘Definitely Include’) and the rest of the components were rated on relatedness (-3 = ‘Highly Unrelated’ to 3 = ‘Highly Related’). For I-CVI (Polit et al., 2007), modified kappas are presented here with the following interpretation: Fair of .40 to .59; Good of .60–.74; and Excellent > .74. Level of agreement (Diaz-Ledezma et al., 2013): Strong (≥80%), Moderate (70-79%), Low (50-69%), and no agreement (<50%

**Supplementary Tables S7:** B) Statistics for Round Two Components Considered for Definition

| Components Considered | Statistics (As stated and [95%CIs]) | Levels of Agreement | Justification | Final Decision |
| --- | --- | --- | --- | --- |
| Disease Activity not captured by DAS28 (hands, shoulders, wrists, elbows and knees) including involvement of other joints (hips, TMJ, feet), other inflammatory features (vasculitis, uveitis, tendonitis or enthesitis) or non-inflammatory features (muscle weakness) | Percentage Include: 76% [61.8%, 86.9%]  Mean Include: 7.48 [6.97, 7.99] | Level: Moderate  I-CVI: 0.76 (Excellent) | Met inclusion criteria, and high means for Rheumatologists and Patients (R2 – Comorbid4). | Definition  Combined as:  Disease Activity not captured by DAS28 (hands, shoulders, wrists, elbows and knees) including involvement of other joints (hips, TMJ, feet), extra-articular manifestations or other inflammatory features (vasculitis, uveitis, tendonitis or enthesitis) or non-inflammatory features (muscle weakness or cachexia) |
| Involvement of other inflammatory features outside of DAS28 e.g. Uveitis or Vasculitis | Percentage Include: 60% [45%, 74%]  Mean Include: 6.62 [6.00, 7.24] | Level: Low  I-CVI = 0.68 (Good) | Crossed threshold for inclusion but already incorporated in above component) (R2 – Comorbid2). |  |
| Presence of Extra-articular Manifestation(s), Complications or associated pathology of Inflammatory Arthritis e.g. Secondary Sjögrens, Nodules, Interstitial Lung Disease, (JIA-associated) Uveitis, Cardiovascular Disease, Anaemia or Chronic leg ulcers | Percentage Include: 62% [47%, 75%]  Mean Include: 6.42 [5.66, 7.18] | Level: Low  I-CVI = 0.71 (Good) | Crossed threshold for inclusion and met criteria for Patients and HCPs (R2 – Comorbid1) – Discussions with Supervisors led to combining above. |  |
| Repeated need of short course steroid tablets or intra-articular injections, that may or may not control flare and localised swelling | Percentage Include: 70% [55.4%, 82.1%]  Mean Include: 6.94 [6.43, 7.45] | Level: Moderate  I-CVI = 0.76 (Excellent) | Met inclusion criteria, and high means for Rheumatologists and Patients (R2 – HCMed2. Free text comment - Any inclusion of steroid use should be very specific about recognition of need to use related to inflammatory arthritis manifestations. | Definition |
| Poor Quality Sleep due to Inflammatory Arthritis | Percentage Include: 60% [45%, 74%]  Mean Include: 6.46 [5.84, 7.08] | Level: Low  I-CVI = 0.44 (Fair) | Percentage and mean value crossed threshold for inclusion. High mean include for Patients. | Model |
| Inability to taper steroid tablets longer term (steroid dependency) | Percentage Include: 58% [43%, 72%]  Mean Include: 6.38 [5.79, 6.97] | Level: Low  I-CVI = 0.54 (Fair) | Percentage crossed threshold for inclusion but mean value did not. | Model |

Please note that components were rated on whether to be included (1 = ‘Definitely Not Include’ to 9 = ‘Definitely Include’) in Round Two. For I-CVI (Polit et al., 2007), modified kappas are presented here with the following interpretation: Fair of .40 to .59; Good of .60–.74; and Excellent > .74. Level of agreement (Diaz-Ledezma et al., 2013): Strong (≥80%), Moderate (70-79%), Low (50-69%), and no agreement (<50%)
