## Supplementary material for "Refractory Inflammatory Arthritis definition and model generated through patient and multi-disciplinary professional modified Delphi process": S8 - Round One Rankings

Supplementary Figure S8: Round One Rankings of Domains by Role Group

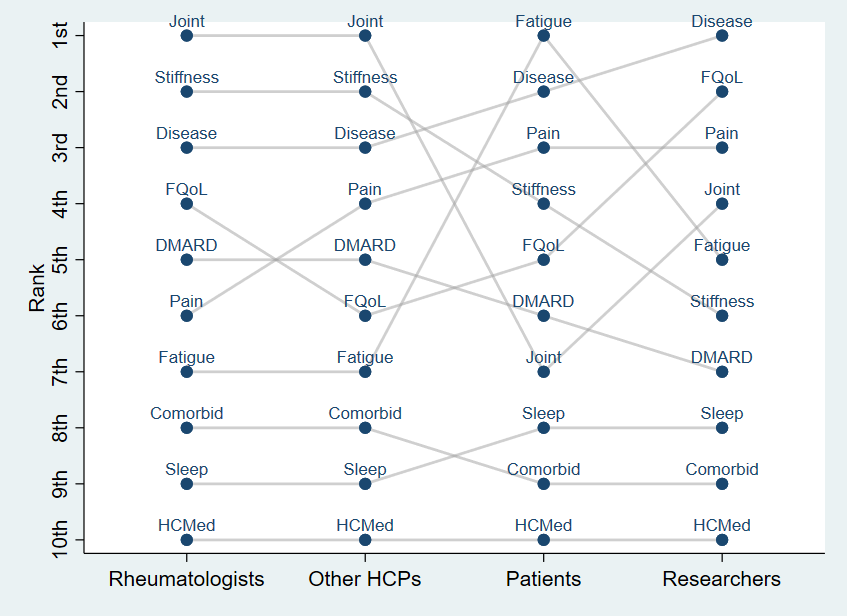

Please note: Joint = Joint Activity, Disease = Disease Activity, FQoL = Functioning and Quality of Life, DMARD = DMARD Experiences, Comorbid = Comorbidities and Wider Involvement Outside of Joints, HCMed = Healthcare and Medications Utilisation
