## Supplementary material for "Refractory Inflammatory Arthritis definition and model generated through patient and multi-disciplinary professional modified Delphi process": S9 - Definition Statistics

**Supplementary Data S9:** Refractory Inflammatory Arthritis Definition Components Statistics

| Domain and Components | Statistics |
| --- | --- |
| 1 - Disease Activity | Percentage Include: 94.87% [82.7%, 99.3%]  Mean Include: 8.28 [7.96, 8.61] |
| Persistently high inflammation and/or symptoms e.g. with or without fluctuations for at least two consecutive clinical visits over the period of at least six months (Combined components) | Mean Related: 2.56 [2.37, 2.76]  Mean diff=0.92, d=0.67  I-CVI = 0.95 (Excellent) |
| Disease Activity not captured by DAS28 (hands, shoulders, wrists, elbows and knees) including involvement of other joints (hips, TMJ, feet), other inflammatory features (vasculitis, uveitis, tendonitis or enthesitis) or non-inflammatory features (muscle weakness or cachexia) | Percentage Include: 76% [61.8%, 86.9%]  Mean Include: 7.48 [6.97, 7.99]  I-CVI = 0.76 (Excellent) |
| Presence of Extra-articular Manifestation(s), Complications or associated pathology of Inflammatory Arthritis e.g. Secondary Sjögrens, Nodules, Interstitial Lung Disease, (JIA-associated) Uveitis, Cardiovascular Disease, Anaemia or Chronic leg ulcers (Combined above) | Percentage Include: 62% [47%, 75%]  Mean Include: 6.42 [5.66, 7.18]  I-CVI = 0.71 (Good) |
| Repeated need of short course steroid tablets or intra-articular injections, that may or may not control flare and localised swelling | Percentage Include: 70% [55.4%, 82.1%]  Mean Include: 6.94 [6.43, 7.45]  I-CVI = 0.76 (Excellent) |
| 2 – Joint Involvement |  |
| 2a - Joint Stiffness | Percentage Include: 92.3% [79.1%, 98.4%]  Mean Include: 8.21 [7.83, 8.58] |
| Joint stiffness during the day (lasting longer than 30-60 minutes in the morning) | Mean Related: 2.54 [2.31, 2.77]  Mean diff=0.18, d=0.18  I-CVI = 0.92 (Excellent) |
| 2b - Joint Activity | Percentage Include: 89.47% [75.2%, 97.1%] Mean Include: 8.21 [7.83, 8.59] |
| One or two persistently active/affected joints despite acceptable control in other joints | Mean Related: 2.00 [1.69, 2.31]  Mean diff=1.11, d=0.78  I-CVI = 0.68 (Good) |
| Accrued damage due to inflammation - Joint erosion(s), deformity(ies) or restrictions in range of movement (may or may not be painful) | Mean Related: 1.37 [0.85, 1.88]  Mean diff=1.68, d=0.95  I-CVI = 0.54 (Fair) |
| 3 - Pain | Percentage Include: 82.1% [66.5%, 92.5%]  Mean Include: 7.79 [7.27, 8.32] |
| Pain in joints e.g. hands and feet | Mean Related: 2.38 [2.13, 2.64]  Mean diff=-0.08, d=-0.10  I-CVI = 0.87 (Excellent) |
| Pain during the day and/or night (Combined components) | Mean Related: 2.10 [1.80, 2.41]  Mean diff=0.03, d=0.03  I-CVI = 0.85 (Excellent) |
| Pain Interference impacting on quality of life (Originally part of Functioning and Quality of Life) | Mean Related: 2.16 [1.81, 2.52]  Mean diff=0.27, d=0.22  I-CVI = 0.76 (Excellent) |
| Domain and Components | Statistics |
| 4 – Fatigue | Percentage Include: 79.5% [63.5%, 90.7%]  Mean Include: 7.54 [6.92, 8.16] |
| Lack of physical energy resulting in difficulties conducting daily activities e.g. washing, dressing | Mean Related: 2.05 [1.70, 2.40]  Mean diff=0.18, d=0.16  I-CVI = 0.74 (Excellent) |
| Lack of mental energy leading to difficulties with concentration and memory | Mean Related: 1.82 [1.45, 2.19]  Mean diff=0.28, d=0.20  I-CVI = 0.63 (Good) |
| 5 - Functioning and Quality of Life | Percentage Include: 78.4% [61.8%, 90.2%]  Mean Include: 7.78 [7.30, 8.26] |
| Problems with self-care e.g. washing/dressing | Mean Related: 2.11 [1.71, 2.51]  Mean diff=0.05, d=0.04  I-CVI = 0.73 (Good) |
| Inability to perform desired activities e.g. hobbies, social, salaried/voluntary work | Mean Related: 2.03 [1.62, 2.43]  Mean diff=0.00, d=0.00  I-CVI = 0.67 (Good) |
| Poor physical function e.g. lack of strength, dexterity, grip | Mean Related: 2.03 [1.64, 2.42]  Mean diff=0.05, d=0.05  I-CVI = 0.73 (Good) |
| Reduced mobility and/or Problems walking, standing or climbing stairs e.g. driving, use of public transport, needing to sit most of the day | Mean Related: 2.00 [1.62, 2.38]  Mean diff=0.00, d=0.00  I-CVI = 0.73 (Good) |
| Disease-related distress e.g. psychological distress related to burden of disease including Physical, Emotional, Social, Treatment/Healthcare Distress | Mean Related: 2.08 [1.72, 2.44]  Mean diff=0.41, d=0.32  I-CVI = 0.70 (Good) |
| 6 – DMARD Experiences | Percentage Include: 73% [55.9%, 86.2%]  Mean Include: 7.41 [6.70,8.11] |
| Primary inefficacy (no response to DMARD at all) and/or Secondary inefficacy (developed ‘resistance’ to DMARD over time) | Mean Related: 2.27 [1.83, 2.71]  Mean diff=1.49, d=0.85  I-CVI = 0.78 (Excellent) |
| Experience of multiple occurrences of inefficacy, intolerability or discontinuation | Mean Related: 2.00 [1.47, 2.53] Mean diff=1.46, d=0.75  I-CVI = 0.76 (Excellent) |

Please note that domains and some components were rated on whether to be included (1 = ‘Definitely Not Include’ to 9 = ‘Definitely Include’) and the rest of the components were rated on relatedness (-3 = ‘Highly Unrelated’ to 3 = ‘Highly Related’). For I-CVI, modified kappas are presented here with the following interpretation: Fair of .40 to .59; Good of .60–.74; and Excellent > .74.

| Components considered for Definition but included in Model | Statistics |
| --- | --- |
| Joint stiffness when resting | Mean Related: 1.69 [1.34, 2.04]  Mean diff=0.05, d=0.04  I-CVI = 0.60 (Good) |
| Joint replacement(s) and/or fusion(s) due to inflammation | Mean Related: 0.79 [0.21, 1.37]  Mean diff=1.26, d=0.68  I-CVI = 0.37 (Poor) |
| Pain in other areas e.g. muscles, neuropathic, regional (e.g. back), widespread etc | Mean Related: 0.85 [0.28, 1.42]  Mean diff=0.36, d=0.20  I-CVI = 0.39 (Poor) |
| Fatigue lasting several hours or all day | Mean Related: 1.85 [1.42, 2.27]  Mean diff=0.05, d=0.04  I-CVI = 0.63 (Good) |
| General Emotional Distress e.g. suspected or diagnosed Depression or Anxiety | Mean Related: 1.65 [1.18, 2.12]  Mean diff=0.65, d=0.44  I-CVI = 0.52 (Fair) |
| Low Self-Efficacy/Esteem/Confidence e.g. perceived low ability to manage and cope with symptoms | Mean Related: 1.27 [0.79, 1.75]  Mean diff=0.54, d=0.36  I-CVI = 0.35 (Poor) |
| Reduced Social Support Network, Relationship breakdowns/difficulties or restriction of social participation | Mean Related: 1.11 [0.55, 1.66]  Mean diff=0.76, d=0.43  I-CVI = 0.36 (Poor) |
| Isolation e.g. quantity of relationships or from unemployment, or Loneliness e.g. perception of disagreement between actual and desired levels of social contact or perceived quality of relationships | Mean Related: 1.03 [0.44, 1.61]  Mean diff=0.73, d=0.40  I-CVI = 0.37 (Poor) |
| Involvement of other inflammatory features outside of DAS28 e.g. Uveitis or Vasculitis (Included above in 1 - Disease Activity) | Percentage Include: 60% [45%, 74%]  Mean Include: 6.62 [6.00, 7.24]  I-CVI = 0.68 (Good) |
| Poor Quality Sleep due to Inflammatory Arthritis | Percentage Include: 60% [45%, 74%]  Mean Include: 6.46 [5.84, 7.08]  I-CVI = 0.44 (Fair) |
| Inability to taper steroid tablets longer term (steroid dependency) | Percentage Include: 58% [43%, 72%]  Mean Include: 6.38 [5.79, 6.97]  I-CVI = 0.54 (Fair) |
