## Supplementary material for "Refractory Inflammatory Arthritis definition and model generated through patient and multi-disciplinary professional modified Delphi process": S10 - Example selection of components

**Supplementary Figure S10:** Example selection of components for inclusion or exclusion from included domain

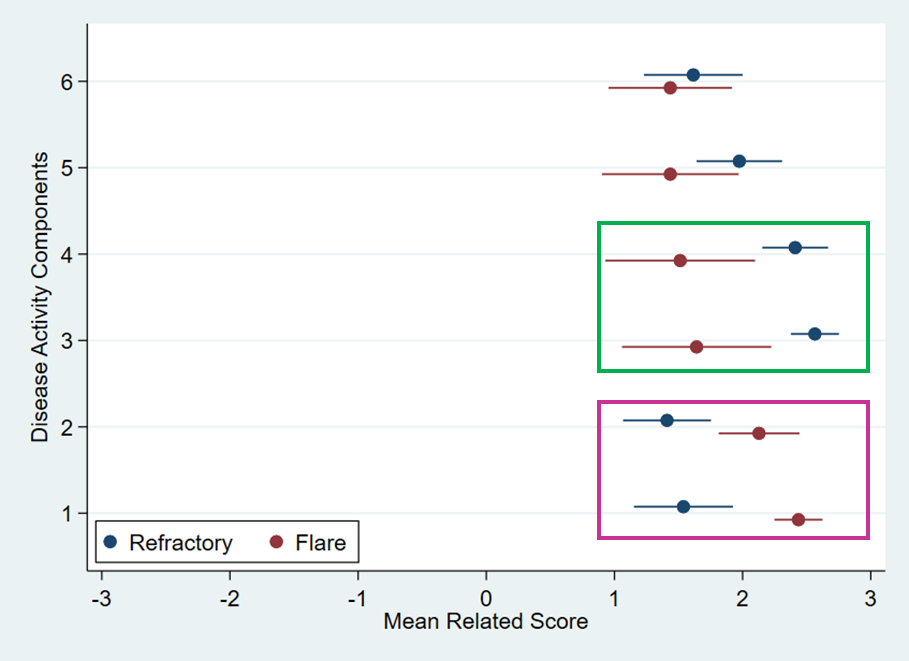

Those highlighted in green have a higher mean related score (2.56 [95%CI 2.37, 2.76]) for RD with a medium-to-large effect size (*d*=0.67) in mean difference (0.92), with Disease Flare and Excellent I-CVI (*k*=0.95) inter-rater agreement and content validity so will be included in the definition. These two refer to “persistently high inflammation and/or symptoms for at least two consecutive clinical visits over the period of at least six months” (have been combined). For example, the two components highlighted in purple in Figure 4 are more related to Disease Flare based on mean related score and mean difference as these are “overall high rating of disease activity/impact from high inflammatory markers or patient report at a single appointment”.
