## Supplementary material for "Refractory Inflammatory Arthritis definition and model generated through patient and multi-disciplinary professional modified Delphi process": S11 - Assessments considered

**Supplementary Table S11**: Assessments initially mapped onto Refractory Inflammatory Arthritis definition

| RIA Definition | | Assessment Methods (including cut-offs) |
| --- | --- | --- |
| Part 1: Treatment | | Treatment History (≥1 csDMARDs, and ≥1 anti-TNF/b/tsDMARDs) (Watts et al., 2013) |
| Part 2: Inflammation | | Abnormal/Raised Inflammatory markers (ESR / CRP based on the local laboratory standards) (Watts et al., 2013)  Physical examination (TJC/SJC) (Watts et al., 2013)  Imaging (Any) (Watts et al., 2013) |
| Part 3: Symptoms and Impact | 1. Disease Activity | Clinical Notes / Consultation / Medical and Treatment History (Watts et al., 2013) DAS28 >3.2 (van der Heijde et al., 1990) or SDAI >11 (Smolen et al., 2003)  RAID (scoring higher on Q5) (Gossec et al., 2011) |
|  | 2. Joint Involvement | (C)HAQ (Symptoms Section) (Fries et al., 1980; Singh et al., 1994)  MSK-HQ (scoring 0-2 on Q1 and/or 2) (Hill et al., 2016)  Consultation / Joint Examination / Clinical Notes (Watts et al., 2013) |
|  | 3. Pain | MSK-HQ (scoring 0-2 on Q1 and/or 2) (Hill et al., 2016)  VAS >3.5 (Boonstra et al., 2014)  EQ5D (scoring 2 or 3 on Q4) (EuroQol Group, 1990)  RAID (scoring higher on Q1) (Gossec et al., 2011) |
|  | 4. Fatigue | BRAF (Higher total score and on items) (Nicklin et al., 2010)  VAS >2.0 (Pollard et al., 2006)  General Fatigue MSK-HQ (scoring 0-2 on Q10) (Hill et al., 2016)  RAID (scoring higher on Q3) (Gossec et al., 2011) |

Please note: DAS28: Disease Activity Score 28 joint count, SDAI: Simplified Disease Activity Index, RAID: Rheumatoid Arthritis Impact of Disease, (C)HAQ: Child or Adult Health Assessment Questionnaire, MSK-HQ: Musculoskeletal Health Questionnaire, VAS: Visual Analogue Scale, EQ5D: EuroQol 5-Dimensions, BRAF: Bristol RA Fatigue Scale

| RIA Definition | | Assessment Methods (including cut-offs) |
| --- | --- | --- |
| Part 3: Symptoms and Impact | 5. Functioning and Quality of Life | (C)HAQ ≥1.5 (Fries et al., 1980; Singh et al., 1994)  EQ5D (scoring 2 or 3 on Q1-3 and Q5) (EuroQol Group, 1990)  WSAS (higher total score and on items 1-4) (Mundt et al., 2002)  MSK-HQ (scoring 0-2 on Q3-7 and Q11) (Hill et al., 2016)  RAID (scoring higher on Q2 and Q6) (Gossec et al., 2011)  RADS (Silke et al., 2021)  Consultation (Watts et al., 2013) |
|  | 6. cs/b/tsDMARD Experiences | Clinical Notes / Treatment and DAS28 History / Consultation (Watts et al., 2013) |

Please note: (C)HAQ: Child or Adult Health Assessment Questionnaire, MSK-HQ: Musculoskeletal Health Questionnaire, EQ5D: EuroQol 5-Dimensions, Work and Social Adjustment Scale, RADS: Rheumatoid Arthritis Distress Scale.
