## Supplementary material for "Refractory Inflammatory Arthritis definition and model generated through patient and multi-disciplinary professional modified Delphi process": S12 - Data from Theoretical Validation

**Supplementary Tables S12:** A) Data capture and synthesis of measures used in the RIA definition across RA/JIA Biologic registries/cohort (Location, Condition and Parts One and Two)

|  |  |  | Part One | Part Two | | |
| --- | --- | --- | --- | --- | --- | --- |
| Name and Reference of Registry or Cohort | Location | Condition | Treatment History | ESR or CRP | T/SJC or A/LJC | Imaging (US or MRI) |
| ABC (Otten et al., 2013) | Netherlands | JIA | ✓ | ✓ | ✓ | ✓ |
| ABioPharm (Barnabe et al., 2018) | Canada | RA | 🗶 | ✓ | ✓ | 🗶 |
| ACTION (Nüßlein et al., 2014) | Worldwide | RA | ✓ | ✓ | ✓ | 🗶 |
| ATTRA (Horák, Skácelová, Hejduk, Smržová, & Pavelka, 2013) | Czech Republic | RA | ✓ | ✓ | ✓ | 🗶 |
| ARAD (Williams, Buchbinder, March, & Lassere, 2011) | Australia | RA & JIA | ✓ | ✓ | ✓ | 🗶 |
| ARAMIS (Singh, 2001) | USA and Canada | RA | ✓ | ✓ | ✓ | 🗶 |
| ARRRA (Karateev et al., 2015) | Russia | RA & JIA | ✓ | ✓ | ✓ | 🗶 |
| ARTIS (Askling et al., 2006) | Sweden | RA | ✓ | ✓ | ✓ | 🗶 |
| BARFOT (Hafström et al., 2019) | Sweden | RA | ✓ | ✓ | ✓ | 🗶 |
| BCRD and BSPAR-ETN (Kearsley-Fleet et al., 2016)  (Hyrich, 2022)* | UK | JIA | ✓ | ✓ | ✓ | 🗶 |
| BIKER (Horneff et al., 2009; Klotsche et al., 2014) | Germany and Austria | RA | ✓ | ✓ | ✓ | 🗶 |
| BIOBADASER (Sanchez-Piedra et al., 2019) | Spain | RA and JIA | ✓ | ✓ | ✓ | 🗶 |
| BIOCURA (Cuppen et al., 2016)  (Hiligsmann, Rademacher, Kaal, Bansback, & Harrison, 2018)* | Netherlands | RA | ✓ | ✓ | ✓ | 🗶 |
| BIOREG (Rintelen et al., 2016) | Austria | RA | ✓ | ✓ | ✓ | 🗶 |
| BioRx.si (Rotar, Hočevar, Rebolj Kodre, Praprotnik, & Tomšič, 2015) | Slovenia | RA | ✓ | ✓ | ✓ | 🗶 |
| BioTRAC (Thorne et al., 2014) | Canada | RA | ✓ | ✓ | ✓ | 🗶 |
| BRASS (Iannaccone et al., 2010) | UK | RA | ✓ | ✓ | ✓ | 🗶 |
| BSR-BR (Hyrich, 2018) | UK | RA | ✓ | ✓ | ✓ | 🗶 |
| CAPS (Adib et al., 2008; Hanns et al., 2016; McErlane et al., 2013) | UK | JIA | ✓ | ✓ | ✓ | 🗶 |
| CARRA (BeukelmanKimura, et al., 2017) | USA and Canada | JIA | ✓ | ✓ | ✓ | ✓ |
| CATCH (Bykerk et al., 2012) | Canada | RA | ✓ | ✓ | ✓ | 🗶 |
| CONAART (Marcos et al., 2010)  (Hiligsmann et al., 2018)* | Argentina | RA | ✓ | ✓ | ✓ | 🗶 |
| CORRONA (Kremer, 2016) | USA | RA | ✓ | ✓ | ✓ | 🗶 |
| DANBIO (Ibfelt, Jensen, & Hetland, 2016) | Denmark | RA | ✓ | ✓ | ✓ | 🗶 |
| DREAM (de Punder et al., 2012)  (Hiligsmann et al., 2018)* | Netherlands | RA | ✓ | ✓ | ✓ | 🗶 |
| ERAS and ERAN, (Young et al., 2011) | UK | RA | Mainly csDMARD | ✓ | ✓ | 🗶 |
| ESPOIR (Combe et al., 2007) | France | RA | ✓ | ✓ | ✓ | ✓ |

Please note: ESR = Erythrocyte Sedimentation Rate, CRP = C-Reactive Protein, T/SJC = Tender/Swollen Joint Count, A/LJC = Active/Limited Joint Count, ABC = Arthritis and Biologicals in Children, ABioPharm = Alberta Biologics Pharmacosurveillance Program, ACTION = Abatacept In Routine Clinical Practice, ATTRA = Czech Biologics Registry, ARAD = Australian Rheumatology Association Database, ARAMIS = Arthritis, Rheumatism and Aging Medical Information System, ARRRA = All-Russian Register of patients with Rheumatoid Arthritis, ARTIS = Swedish National Biologics Registry, BARFOT = Better Anti-Rheumatic Pharmacotherapy, BCRD = Biologics for Children with Rheumatic, Diseases, BSPAR-ETN = British Society for Paediatric and Adolescent Rheumatology – Etanercept, BIKER = German Biologics JIA Registry, BIOBADASER = Spanish registry of adverse events involving biological therapies in rheumatic diseases, BIOCURA = Biologicals and Outcome Compared and Predicted in Utrecht Region in Rheumatoid Arthritis study, BIOREG = Austrian Biologics Registry for inflammatory rheumatic disease, BioRx.si, = Slovenian prospective national on-line biologics registry, BioTRAC = Biologic Treatment Registry Across Canada, BRASS = Brigham and Women's Hospital Rheumatoid Arthritis Sequential Study, BSR-BR = British Society of Rheumatology Biologics Register, CAPS = Childhood Arthritis Prospective Study, CARRA = Childhood Arthritis and Rheumatology Research Alliance, CATCH = Canadian Early Arthritis Cohort, CONAART = Argentine Consortium for Early Arthritis, CORRONA = Consortium of Rheumatology Researchers of North America, DANBIO = Danish biologics register for rheumatologic diseases, DREAM = Dutch Rheumatoid Arthritis Monitoring registry, ERAS/N = Early Rheumatoid Arthritis Study/Network, ESPOIR = Etude et Suivi des Polyarthrites Indifférenciées Récentes.

|  |  |  | Part One | Part Two | | |
| --- | --- | --- | --- | --- | --- | --- |
| Name and Reference of Registry or Cohort | Location | Condition | Treatment History | ESR or CRP | T/SJC or A/LJC | Imaging (US or MRI) |
| GISEA (Lapadula, Ferraccioli, Ferri, Punzi, & Trotta, 2011) | Italy | RA | ✓ | ✓ | ✓ | Not clear |
| GLADAR (Massardo et al., 2012) | Latin America | RA | 🗶 | ✓ | ✓ | 🗶 |
| ICON (Sengler et al., 2015) | Germany | JIA | ✓ | ✓ | ✓ | 🗶 |
| IMPARTS (Matcham, Norton, Steer, & Hotopf, 2016; Rayner et al., 2014) | UK | RA | 🗶 | ✓ | ✓ | 🗶 |
| IORRA (Yamanaka et al., 2007)  (Hiligsmann et al., 2018)* | Japan | RA | ✓ | ✓ | ✓ | 🗶 |
| JACS (Beukelman, Anink, et al., 2017) | Australia | JIA | Not Clear | ✓ | ✓ | 🗶 |
| JUMBO (Minden et al., 2012) | Germany | JIA | ✓ | ✓ | ✓ | 🗶 |
| METEOR (van den Berg, van der Heijde, Landewé, van Lambalgen, & Huizinga, 2014) | Worldwide | RA | ✓ | ✓ | ✓ | 🗶 |
| NDB (Zink, Listing, Klindworth, & Zeidler, 2001)  (Hiligsmann et al., 2018)* | Germany | RA | ✓ | ✓ | ✓ | 🗶 |
| NDB (Wolfe & Michaud, 2011) | USA | RA | ✓ | ✓ | ✓ | 🗶 |
| NEIAA (British Society of Rheumatology, 2019) | UK | RA & JIA | Some | ✓ | ✓ | 🗶 |
| Nijmegen Early RA Cohort (Welsing & van Riel, 2004)  (Hiligsmann et al., 2018)* | Netherlands | RA | ✓ | ✓ | ✓ | 🗶 |
| NoAR (Druce, Jones, Macfarlane, Verstappen, & Basu, 2015; Symmons & Silman, 2003) | UK | RA | Mainly csDMARD | ✓ | ✓ | 🗶 |
| Nordic JIA Cohort (Nordal et al., 2011) | Sweden, Finland, Denmark, Norway, Iceland | JIA | ✓ | ✓ | ✓ | 🗶 |
| NoRDMARD (Olsen, Haavardsholm, Moholt, Kvien, & Lie, 2014) | Norway | RA & JIA | ✓ | ✓ | ✓ | 🗶 |
| NPRD (Minden, Niewerth, Listing, & Zink, 2002) | Germany | JIA | Mainly csDMARD | Not clear | ✓ | 🗶 |
| ORAR (Kvien & Uhlig, 2004) | Norway | RA | ✓ | ✓ | ✓ | 🗶 |
| PHARMACHILD (Beukelman, Anink, et al., 2017) | Worldwide | JIA | Not Clear | ✓ | ✓ | Not clear |
| QUEST-RA (Sokka et al., 2007) | Worldwide | RA | ✓ | ✓ | ✓ | 🗶 |
| RABBIT (Gerhold et al., 2015) | Germany | RA | ✓ | ✓ | ✓ | 🗶 |
| RADIUS1 (Gibofsky et al., 2011) | USA | RA | ✓ | ✓ | ✓ | 🗶 |
| ReACCh Out (Oen et al., 2009) | Canada | JIA | ✓ | ✓ | ✓ | 🗶 |
| REACH (Alves et al., 2011; Kuijper et al., 2014) | Netherlands | RA | ✓ | ✓ | ✓ | 🗶 |
| REUMA.PT (Canhão, Faustino, Martins, & Fonseca, 2011) | Portugal | RA & JIA | ✓ | ✓ | ✓ | 🗶 |
| Rhumadata© (Choquette et al., 2019) | Canada | RA | ✓ | ✓ | ✓ | 🗶 |

Please note: ESR = Erythrocyte Sedimentation Rate, CRP = C-Reactive Protein, T/SJC = Tender/Swollen Joint Count, A/LJC = Active/Limited Joint Count, GISEA = Italian Group for the Study of Early Arthritis, GLADAR = Grupo Latino Americano de Estudio de Artritis Reumatoide, ICON = Inception Cohort of Newly diagNosed patients with JIA,, IMPARTS = Integrating Mental & Physical healthcare: Research, Training & Services, IORRA = Institute of Rheumatology, Rheumatoid Arthritis, JACS = Juvenile arthritis cohort study, JUMBO = Juvenile arthritis MTX/Biologics long-term Observation, METEOR = Measurement of Efficacy of Treatment in the “Era of Outcome” in Rheumatology, NDB = national database of the German Collaborative Arthritis Centres, NDB = National Data Bank for Rheumatic Diseases, NEIAA = National Early Inflammatory Arthritis Audit, NoAR = Norfolk Arthritis Register, NoRDDMARD = Norwegian Antirheumatic Drug Register, NPRD = National Paediatric Rheumatology Database, ORAR = Oslo Rheumatoid Arthritis Register, PHARMACHILD = Pharmacovigilance in JIA patients treated with biologic agents and/or MTX, QUEST-RA = Quantitative Patient Questionnaires in Standard Monitoring of Patients with Rheumatoid Arthritis, RABBIT = German register for long-term observation of biologics in RA, RADIUS1 = Rheumatoid Arthritis Disease-Modifying Antirheumatic Drug Intervention and Utilization Study Cohort1, ReACCh Out = Research in Arthritis in Canadian Children emphasizing Outcomes, REACH = Rotterdam Early Arthritis Cohort, REUMA.PT = Rheumatic Diseases Portuguese Register.

|  |  |  | Part One | Part Two | | |
| --- | --- | --- | --- | --- | --- | --- |
| Name and Reference of Registry or Cohort | Location | Condition | Treatment History | ESR or CRP | T/SJC or A/LJC | Imaging (US or MRI) |
| ROB-FIN (Nordström et al., 2006) | Finland | RA | ✓ | ✓ | ✓ | 🗶 |
| RRBR (Codreanu, Mogosan, Ionescu, Ancuta, & Opris, 2014) | Romania | RA | ✓ | ✓ | ✓ | ✓ |
| SCQM (Uitz, Fransen, Langenegger, & Stucki, 2000) | Switzerland | RA | ✓ | ✓ | ✓ | 🗶 |
| SRQ (Eriksson, Askling, & Arkema, 2014)  (Hiligsmann et al., 2018)* | Sweden | RA | ✓ | ✓ | ✓ | 🗶 |
| STURE (Askling et al., 2006) | Sweden | RA | ✓ | ✓ | ✓ | 🗶 |
| Swedish JIA Register (Beukelman, Anink, et al., 2017) | Sweden | JIA | Not Clear | ✓ | ✓ | 🗶 |
| TURKBIO (Önen et al., 2022) | Turkey | RA | ✓ | ✓ | ✓ | 🗶 |
| VARAR (Mikuls et al., 2010) | USA | RA | ✓ | ✓ | ✓ | 🗶 |
| WC (Kovalchik, Charles-Schoeman, Khanna, & Paulus, 2012) | USA and Mexico | RA | Mainly csDMARD | ✓ | ✓ | 🗶 |

Please note: ESR = Erythrocyte Sedimentation Rate, CRP = C-Reactive Protein, T/SJC = Tender/Swollen Joint Count, A/LJC = Active/Limited Joint Count, ROB-FIN = National Register of Biological Treatment in Finland, RRBR = Romanian Registry of Rheumatic Diseases, SCQM = Swiss Clinical Quality Management in Rheumatic Diseases, SRQ = Swedish Rheumatology Quality Registries, STURE = Stockholm TNF follow-Up Registry, TURKBIO = Turkish Biologics Register, VARAR = Veterans Affairs Rheumatoid Arthritis Registry, WC = Western Consortium of Practicing Rheumatologists.

**Supplementary Tables S12:** B) Data capture and synthesis of measures used in the RIA definition across RA/JIA Biologic registries/cohort (Part Three)

|  | Part Three | | | | | | | | | | | | | | | | | |
| --- | --- | --- | --- | --- | --- | --- | --- | --- | --- | --- | --- | --- | --- | --- | --- | --- | --- | --- |
| Name and Reference of Registry or Cohort | Treatment History | Clinical Notes | DAS28/JADAS or C/SDAI | RAID | Manifestations/ Features | Steroid Use | Stiffness | T/SJC or A/LJC | Imaging (Radiographs) | MSK-HQ | Pain VAS | EQ-5D | BRAF | Fatigue VAS | (C)HAQ | WSAS | RADS | Treatment Change Reasons |
| ABC  (Otten et al., 2013) | ✓ | ✓ | 🗶 | 🗶 | 🗶 | ✓ | 🗶 | ✓ | 🗶 | 🗶 | 🗶 (PtVAS) | 🗶 | 🗶 | 🗶 (PtVAS) | ✓ | 🗶 | 🗶 | ✓ |
| ABioPharm (Barnabe et al., 2018) | 🗶 | 🗶 | ✓ | 🗶 | 🗶 | 🗶 | ✓ | ✓ | 🗶 | 🗶 | ✓ | ✓ | 🗶 (SF-36) | ✓ | ✓ | 🗶 (study-specific) | 🗶 (SF-36) | 🗶 |
| ACTION (Nüßlein et al., 2014) | ✓ | ✓ | ✓ | 🗶 | 🗶 | ✓ | 🗶 | ✓ | ✓ | 🗶 | ✓ | 🗶 | 🗶 | 🗶 (PtVAS) | ✓ | 🗶 | 🗶 | ✓ |
| ATTRA  (Horák et al., 2013) | ✓ | ✓ | ✓ | 🗶 | 🗶 | ✓ | 🗶 | ✓ | 🗶 | 🗶 (SF-36) | 🗶 (PtVAS) | ✓ | 🗶 (SF-36) | 🗶 (SF-36) | ✓ | 🗶 (SF-36) | 🗶 (SF-36) | ✓ |
| ARAD  (Williams et al., 2011) | ✓ | ✓ | ✓ | ✓ | 🗶 | ✓ | 🗶 | ✓ | 🗶 | 🗶 (AQoL & PEDs-QL) | 🗶 (PtVAS) | ✓ | 🗶 (SF-36) | 🗶 (PtVAS) | ✓ | 🗶 (AQoL and PEDs-QL) | 🗶 (SF-36) | ✓ |
| ARAMIS (Singh, 2001) | ✓ | ✓ | ✓ | 🗶 | 🗶 | ✓ | 🗶 | ✓ | 🗶 | 🗶 | ✓ | 🗶 | 🗶 (SF-36) | 🗶 (SF-36) | ✓ | 🗶 (SF-36) | 🗶 (AIMS-DA & SF-36) | ✓ |
| ARRRA  (Karateev et al., 2015) | ✓ | ✓ | ✓ | 🗶 (RAPID3) | 🗶 | ✓ | 🗶 | ✓ | 🗶 | 🗶 (RAPID3) | 🗶 (RAPID3 & PtVAS) | ✓ | 🗶 | 🗶 (PtVAS) | ✓ | 🗶 | 🗶 | 🗶 |
| ARTIS (Askling et al., 2006) | ✓ | ✓ | ✓ | 🗶 | 🗶 | ✓ | 🗶 | ✓ | 🗶 | 🗶 | ✓ | 🗶 | 🗶 | 🗶 (PtVAS) | ✓ | 🗶 | 🗶 | Not clear |

Please note: T/SJC = Tender/Swollen Joint Count, A/LJC = Active/Limited Joint Count, DAS28 = Disease Activity Score-28 Joint count, JADAS = Juvenile Arthritis Disease Activity Score, C/SDAI = Clinical/Simplified Disease Activity Index, RAID = Rheumatoid Arthritis Impact of Disease, (C)HAQ = (Child) Health Assessment Questionnaire , MSK-HQ = Musculoskeletal Health Questionnaire, VAS = Visual Analogue Scale, EQ-5D = EuroQol 5 Dimensions, BRAF = Bristol Rheumatoid Arthritis Fatigue, WSAS = Work and Social Adjustment Scale, RADS = Rheumatoid Arthritis Distress Scale, ABC = Arthritis and Biologicals in Children, ABioPharm = Alberta Biologics Pharmacosurveillance Program, ACTION = Abatacept In Routine Clinical Practice, ATTRA = Czech Biologics Registry, ARAD = Australian Rheumatology Association Database, ARAMIS = Arthritis, Rheumatism and Aging Medical Information System, ARRRA = All-Russian Register of patients with Rheumatoid Arthritis, ARTIS = Swedish National Biologics Registry, RAPID3 = Routine Assessment of Patient Index Data 3, PtVAS = Patient Global VAS, SF-36 = Short Form – 36 Item, AQoL = Assessment of Quality of Life, Peds-QL = Pediatric Quality of Life Inventory, AIMS-DA = Arthritis Impact Measurement Scales depression and anxiety scales

|  | Part Three | | | | | | | | | | | | | | | | | |
| --- | --- | --- | --- | --- | --- | --- | --- | --- | --- | --- | --- | --- | --- | --- | --- | --- | --- | --- |
| Name and Reference of Registry or Cohort | Treatment History | Clinical Notes | DAS28/JADAS or C/SDAI | RAID | Manifestations/ Features | Steroid Use | Stiffness | T/SJC or A/LJC | Imaging (Radiographs) | MSK-HQ | Pain VAS | EQ-5D | BRAF | Fatigue VAS | (C)HAQ | WSAS | RADS | Treatment Change Reasons |
| BARFOT (Hafström et al., 2019) | ✓ | ✓ | ✓ | 🗶 | 🗶 | ✓ | 🗶 | ✓ | ✓ | 🗶 | ✓ | 🗶 | 🗶 | 🗶 (PtVAS) | ✓ | 🗶 | 🗶 | Not clear |
| BCRD and BSPAR-ETN (Kearsley-Fleet et al., 2016)  (Hyrich, 2022)* | ✓ | ✓ | ✓ | 🗶 (CHU-9D)* | 🗶 | ✓ | 🗶 | ✓ | 🗶 | 🗶 (CHU-9D)* | ✓ | 🗶 (EQ-5D-Y)* | 🗶 | 🗶 (PtVAS & CHU-9D)* | ✓ | 🗶 (CHU-9D)* | 🗶 (CHU-9D)* | ✓ |
| BIKER (Horneff et al., 2009; Klotsche et al., 2014) | ✓ | ✓ | 🗶 (PedACR) | 🗶 (PedsQL) | 🗶 | ✓ | ✓ | ✓ | 🗶 | 🗶 (PedsQL) | 🗶 (PtVAS) | 🗶 | 🗶 | 🗶 (PtVAS) | ✓ | 🗶 (PedsQL) | 🗶 (PedsQL) | ✓ |
| BIOBADASER (Sanchez-Piedra et al., 2019) | ✓ | ✓ | ✓ | 🗶 | 🗶 | Not clear | 🗶 | ✓ | 🗶 | 🗶 | 🗶 (PtVAS) | 🗶 | 🗶 | 🗶 (PtVAS) | 🗶 | 🗶 | 🗶 | ✓ |
| BIOCURA (Cuppen et al., 2016)  (Hiligsmann et al., 2018)* | ✓ | ✓ | ✓ | 🗶 | 🗶 | ✓ | 🗶 | ✓ | 🗶 | 🗶 | ✓ | ✓ | 🗶 (SF-36)* | 🗶 (SF-36)* | ✓ | 🗶 (SF-36)* | 🗶 (SF-36)* | Some |
| BIOREG (Rintelen et al., 2016) | ✓ | ✓ | ✓ | 🗶 | 🗶 | ✓ | 🗶 | ✓ | 🗶 | 🗶 | 🗶 (RADAI-5) | 🗶 | 🗶 | 🗶 (PtVAS) | ✓ | 🗶 | 🗶 | ✓ |
| BioRx.si (Rotar et al., 2015) | ✓ | ✓ | ✓ | 🗶 | 🗶 | ✓ | 🗶 | ✓ | 🗶 | 🗶 | ✓ | 🗶 | 🗶 | 🗶 (PtVAS) | ✓ | 🗶 | 🗶 | 🗶 |
| BioTRAC (Thorne et al., 2014) | ✓ | ✓ | ✓ | 🗶 | 🗶 | 🗶 | ✓ | ✓ | 🗶 | 🗶 | ✓ | 🗶 | 🗶 | 🗶 (PtVAS) | ✓ | 🗶 | 🗶 | ✓ |

Please Note: T/SJC = Tender/Swollen Joint Count, A/LJC = Active/Limited Joint Count, DAS28 = Disease Activity Score-28 Joint count, JADAS = Juvenile Arthritis Disease Activity Score, C/SDAI = Clinical/Simplified Disease Activity Index,, RAID = Rheumatoid Arthritis Impact of Disease, (C)HAQ = (Child) Health Assessment Questionnaire , MSK-HQ = Musculoskeletal Health Questionnaire, VAS = Visual Analogue Scale, EQ-5D = EuroQol 5 Dimensions, BRAF = Bristol Rheumatoid Arthritis Fatigue, WSAS = Work and Social Adjustment Scale, RADS = Rheumatoid Arthritis Distress Scale, BARFOT = Better Anti-Rheumatic Pharmacotherapy, BCRD = Biologics for Children with Rheumatic, Diseases, BSPAR-ETN = British Society for Paediatric and Adolescent Rheumatology – Etanercept, BIKER = German Biologics JIA Registry, BIOBADASER = Spanish registry of adverse events involving biological therapies in rheumatic diseases, BIOCURA = Biologicals and Outcome Compared and Predicted in Utrecht Region in Rheumatoid Arthritis study, BIOREG = Austrian Biologics Registry for inflammatory rheumatic disease, BioRx.si, = Slovenian prospective national on-line biologics registry, BioTRAC = Biologic Treatment Registry Across Canada, PtVAS = Patient Global VAS, CHU-9D = Child Health Utility Index – 9 item, EQ-5D-Y = EuroQol 5 Dimensions Youth, PedACR = ACR paediatric response measure, Peds-QL = Pediatric Quality of Life Inventory, SF-36 = Short Form – 36 Item, RADAI(5) = Rheumatoid Arthritis Disease Activity Index (5 item) *Collected but data Not currently published

|  | Part Three | | | | | | | | | | | | | | | | | |
| --- | --- | --- | --- | --- | --- | --- | --- | --- | --- | --- | --- | --- | --- | --- | --- | --- | --- | --- |
| Name and Reference of Registry or Cohort | Treatment History | Clinical Notes | DAS28/JADAS or C/SDAI | RAID | Manifestations/ Features | Steroid Use | Stiffness | T/SJC or A/LJC | Imaging (Radiographs) | MSK-HQ | Pain VAS | EQ-5D | BRAF | Fatigue VAS | (C)HAQ | WSAS | RADS | Treatment Change Reasons |
| BRASS (Iannaccone et al., 2010) | ✓ | ✓ | ✓ | 🗶 | ✓ | ✓ | ✓ | ✓ | 🗶 | 🗶 (ASES & SF-12) | ✓ | ✓ | 🗶 | Not clear | ✓ | 🗶 (Social Support) | 🗶 (SF-12, MHI & PHQ-9) | ✓ |
| BSR-BR (Hyrich, 2018) | ✓ | ✓ | ✓ | 🗶 | ✓ | ✓ | 🗶 | ✓ | 🗶 | 🗶 (SF-36 & B-IPQ) | 🗶 (PtVAS) | ✓ | 🗶 (SF-36) | 🗶 (SF-36) | ✓ | 🗶 (WPS) | 🗶 (SF-36 & B-IPQ) | ✓ |
| CAPS (Adib et al., 2008; Hanns et al., 2016; McErlane et al., 2013) | ✓ | ✓ | ✓ | 🗶 | ✓ | ✓ | 🗶 | ✓ | 🗶 | 🗶 | ✓ | 🗶 | 🗶 | 🗶 (PtVAS) | ✓ | 🗶 | 🗶 (MFQ) | 🗶 |
| CARRA (BeukelmanKimura, et al., 2017) | ✓ | ✓ | ✓ | 🗶 (PGH-7) | ✓ | ✓ | ✓ | ✓ | ✓ | 🗶 (PGH-7) | ✓ | 🗶 | 🗶 | 🗶 (PtVAS) | ✓ | 🗶 (PGH-7) | 🗶 (PGH-7) | ✓ |
| CATCH (Bykerk et al., 2012) | ✓ | ✓ | 🗶 (RADAI) | 🗶 | ✓ | ✓ | ✓ | ✓ | ✓ | 🗶 | 🗶 (PtVAS) | 🗶 | 🗶 | 🗶 (PtVAS) | ✓ | 🗶 | 🗶 | 🗶 |
| CONAART (Marcos et al., 2010)  (Hiligsmann et al., 2018)* | ✓ | ✓ | ✓ | 🗶 (RAPID3)* | ✓ | 🗶 | ✓ | ✓ | ✓ | 🗶 (RAQoL & RAPID3)* | 🗶 (PtVAS & RAPID3)* | ✓ | 🗶 | 🗶 (PtVAS) | ✓ | 🗶 | 🗶 | 🗶 |
| CORRONA (Kremer, 2016) | ✓ | ✓ | ✓ | 🗶 | 🗶 | 🗶 | 🗶 | ✓ | ✓ | 🗶 | 🗶 (PtVAS) | ✓ | 🗶 | 🗶 (PtVAS) | ✓ | 🗶 | 🗶 | ✓ |

Please Note: T/SJC = Tender/Swollen Joint Count, A/LJC = Active/Limited Joint Count, DAS28 = Disease Activity Score-28 Joint count, JADAS = Juvenile Arthritis Disease Activity Score, C/SDAI = Clinical/Simplified Disease Activity Index, RAID = Rheumatoid Arthritis Impact of Disease, (C)HAQ = (Child) Health Assessment Questionnaire , MSK-HQ = Musculoskeletal Health Questionnaire, VAS = Visual Analogue Scale, EQ-5D = EuroQol 5 Dimensions, BRAF = Bristol Rheumatoid Arthritis Fatigue, WSAS = Work and Social Adjustment Scale, RADS = Rheumatoid Arthritis Distress Scale, BRASS = Brigham and Women's Hospital Rheumatoid Arthritis Sequential Study, BSR-BR = British Society of Rheumatology Biologics Register, CAPS = Childhood Arthritis Prospective Study, CARRA = Childhood Arthritis and Rheumatology Research Alliance, CATCH = Canadian Early Arthritis Cohort, CONAART = Argentine Consortium for Early Arthritis, CORRONA = Consortium of Rheumatology Researchers of North America, PtVAS = Patient Global VAS, SF-12/36 = Short Form – 12/36 Item, ASES = Arthritis Self-Efficacy Scale, MHI = Mental Health Inventory, PHQ-9 = Patient Health Questionnaire 9-items, B-IPQ = Brief Illness Perceptions Questionnaire, WPS = Work Productivity Survey, MFQ = Mood and Feelings Questionnaire, PGH-7 = Pediatric Global Health 7-item, RADAI = Rheumatoid Arthritis Disease Activity Index, RAPID3 = Routine Assessment of Patient Index Data 3, RAQoL = RA Quality of Life, *Collected but data Not currently published

|  | Part Three | | | | | | | | | | | | | | | | | |
| --- | --- | --- | --- | --- | --- | --- | --- | --- | --- | --- | --- | --- | --- | --- | --- | --- | --- | --- |
| Name and Reference of Registry or Cohort | Treatment History | Clinical Notes | DAS28/JADAS or C/SDAI | RAID | Manifestations/ Features | Steroid Use | Stiffness | T/SJC or A/LJC | Imaging (Radiographs) | MSK-HQ | Pain VAS | EQ-5D | BRAF | Fatigue VAS | (C)HAQ | WSAS | RADS | Treatment Change Reasons |
| DANBIO (Ibfelt et al., 2016) | ✓ | ✓ | ✓ | 🗶 | 🗶 | ✓ | 🗶 | ✓ | ✓ | 🗶 | ✓ | ✓ | 🗶 | ✓ | ✓ | 🗶 | 🗶 | ✓ |
| DREAM (de Punder et al., 2012)  (Hiligsmann et al., 2018)* | ✓ | ✓ | ✓ | 🗶 | 🗶 | ✓ | 🗶 | ✓ | 🗶 | 🗶 | 🗶 (PtVAS) | 🗶 | 🗶 | 🗶 (PtVAS) | ✓ | 🗶 | 🗶  (SF-6D)* | 🗶 |
| ERAS and ERAN, (Young et al., 2011) | Mainly csDMARD | ✓ | ✓ | 🗶 (SF-36) | ✓ | ✓ | 🗶 | ✓ | ✓ | 🗶 (SF-36) | 🗶 (PtVAS) | 🗶 (SF-36) | 🗶 (SF-36) | 🗶 (SF-36) | ✓ | 🗶 (SF-36) | 🗶 (SF-36) | ✓ |
| ESPOIR (Combe et al., 2007) | ✓ | ✓ | ✓ | 🗶 (AIMS2) | 🗶 | 🗶 | 🗶 | ✓ | ✓ | 🗶 (AIMS2) | 🗶 (PtVAS & AIMS2) | ✓ | 🗶 (SF-36) | 🗶 (SF-36) | ✓ | 🗶 (AIMS2 & SF-36) | 🗶 (SF-36) | 🗶 |
| GISEA (Lapadula et al., 2011) | ✓ | ✓ | ✓ | 🗶 | ✓ | ✓ | 🗶 | ✓ | ✓ | 🗶 | 🗶 (PtVAS) | ✓ | 🗶 | 🗶 (PtVAS) | ✓ | 🗶 (work time lost) | 🗶 | ✓ |
| GLADAR (Massardo et al., 2012) | 🗶 | 🗶 | ✓ | 🗶 | 🗶 | 🗶 | 🗶 | ✓ | ✓ | 🗶 | ✓ | 🗶 | 🗶 | 🗶 (PtVAS) | ✓ | 🗶 | 🗶 | 🗶 |
| ICON (Sengler et al., 2015) | ✓ | ✓ | ✓ | 🗶 (PedsQL) | ✓ | ✓ | ✓ | ✓ | 🗶 | 🗶 (PedsQL) | ✓ | 🗶 (PedsQL) | 🗶 | 🗶 (PtVAS) | ✓ | 🗶 | 🗶 | 🗶 |
| IMPARTS (Matcham et al., 2016; Rayner et al., 2014) | 🗶 | ✓ | ✓ | 🗶 (SF-36) | 🗶 | 🗶 | 🗶 | ✓ | 🗶 | 🗶 (SF-36) | ✓ | 🗶 | 🗶 (SF-36) | ✓ | ✓ | 🗶 (SF-36) | 🗶 (PHQ9, GAD7 & SF-36) | 🗶 |

Please Note: T/SJC = Tender/Swollen Joint Count, A/LJC = Active/Limited Joint Count, DAS28 = Disease Activity Score-28 Joint count, JADAS = Juvenile Arthritis Disease Activity Score, C/SDAI = Clinical/Simplified Disease Activity Index, RAID = Rheumatoid Arthritis Impact of Disease, (C)HAQ = (Child) Health Assessment Questionnaire , MSK-HQ = Musculoskeletal Health Questionnaire, VAS = Visual Analogue Scale, EQ-5D = EuroQol 5 Dimensions, BRAF = Bristol Rheumatoid Arthritis Fatigue, WSAS = Work and Social Adjustment Scale, RADS = Rheumatoid Arthritis Distress Scale, DANBIO = Danish biologics register for rheumatologic diseases, DREAM = Dutch Rheumatoid Arthritis Monitoring registry, ERAS/N = Early Rheumatoid Arthritis Study/Network, ESPOIR = Etude et Suivi des Polyarthrites Indifférenciées Récentes, GISEA = Italian Group for the Study of Early Arthritis, GLADAR = Grupo Latino Americano de Estudio de Artritis Reumatoide, ICON = Inception Cohort of Newly diagnosed patients with JIA, IMPARTS = Integrating Mental & Physical healthcare: Research, Training & Services, PtVAS = Patient Global VAS, SF-6D = Short Form – 6 Dimensions, SF-36 = Short Form – 36 Item. AIMS2 = Arthritis Impact Measurement Scales 2, Peds-QL = Pediatric Quality of Life Inventory, PHQ9 = Patient Health Questionnaire 9-item, GAD7 = Generalised Anxiety Disorder 7-item, *Collected but data Not currently published

|  | Part Three | | | | | | | | | | | | | | | | | |
| --- | --- | --- | --- | --- | --- | --- | --- | --- | --- | --- | --- | --- | --- | --- | --- | --- | --- | --- |
| Name and Reference of Registry or Cohort | Treatment History | Clinical Notes | DAS28/JADAS or C/SDAI | RAID | Manifestations/ Features | Steroid Use | Stiffness | T/SJC or A/LJC | Imaging (Radiographs) | MSK-HQ | Pain VAS | EQ-5D | BRAF | Fatigue VAS | (C)HAQ | WSAS | RADS | Treatment Change Reasons |
| IORRA (Yamanaka et al., 2007)  (Hiligsmann et al., 2018)* | ✓ | ✓ | ✓ | 🗶 | 🗶 | ✓ | 🗶 | ✓ | 🗶 | 🗶 | ✓ | ✓ | 🗶 | 🗶 | ✓ | 🗶 | 🗶 | 🗶 |
| JACS (Beukelman, Anink, et al., 2017) | Not Clear | Not Clear | ✓ | 🗶 | ✓ | Not Clear | Not Clear | ✓ | 🗶 | 🗶 | ✓ | 🗶 | 🗶 | 🗶 (PtVAS) | ✓ | 🗶 | 🗶 | Not Clear |
| JUMBO (Minden et al., 2012) | ✓ | ✓ | 🗶 (PhVAS) | 🗶 | ✓ | ✓ | ✓ | ✓ | 🗶 | 🗶 (PtVAS) | ✓ | 🗶 | 🗶 (PtVAS) | ✓ | ✓ | 🗶 (PtVAS) | 🗶 (PtVAS) | ✓ |
| METEOR (van den Berg et al., 2014) | ✓ | ✓ | ✓ | 🗶 (RAPID3) | 🗶 | ✓ | 🗶 | ✓ | 🗶 | 🗶 (RAPID3) | ✓ | 🗶 | 🗶 | 🗶 (PtVAS) | ✓ | 🗶 | 🗶 | ✓ |
| NDB (Zink et al., 2001)  (Hiligsmann et al., 2018)* | ✓ | ✓ | ✓ | ✓ | 🗶 | ✓ | 🗶 | ✓ | 🗶 | 🗶 | ✓ | ✓ | 🗶 | 🗶 (PtVAS) | ✓ | 🗶 (study specific) | 🗶 | 🗶 |
| NDB (Wolfe & Michaud, 2011) | ✓ | ✓ | 🗶 (RA Activity & RADAI) | 🗶 | 🗶 | ✓ | ✓ | ✓ | 🗶 | 🗶 (PtVAS) | ✓ | ✓ | 🗶 (PtVAS) | ✓ | ✓ | 🗶 (PtVAS) | 🗶 (PtVAS) | ✓ |
| NEIAA (British Society of Rheumatology, 2019) | Some | ✓ | ✓ | 🗶 | 🗶 | ✓ | 🗶 (MSK-HQ) | ✓ | 🗶 | ✓ | 🗶 (MSK-HQ) | 🗶 | 🗶 | 🗶 (MSK-HQ) | ✓ | 🗶 (WPAI) | 🗶 (PHQ4ADS) | 🗶 |

Please Note: ESR = Erythrocyte Sedimentation Rate, CRP = C-Reactive Protein, T/SJC = Tender/Swollen Joint Count, A/LJC = Active/Limited Joint Count, DAS28 = Disease Activity Score-28 Joint count, JADAS = Juvenile Arthritis Disease Activity Score, C/SDAI = Clinical/Simplified Disease Activity Index, RAID = Rheumatoid Arthritis Impact of Disease, (C)HAQ = (Child) Health Assessment Questionnaire , MSK-HQ = Musculoskeletal Health Questionnaire, VAS = Visual Analogue Scale, EQ-5D = EuroQol 5 Dimensions, BRAF = Bristol Rheumatoid Arthritis Fatigue, WSAS = Work and Social Adjustment Scale, RADS = Rheumatoid Arthritis Distress Scale, IORRA = Institute of Rheumatology, Rheumatoid Arthritis, JACS = Juvenile arthritis cohort study, JUMBO = Juvenile arthritis MTX/Biologics long-term Observation, METEOR = Measurement of Efficacy of Treatment in the “Era of Outcome” in Rheumatology, NDB = national database of the German Collaborative Arthritis Centres, NDB = National Data Bank for Rheumatic Diseases, NEIAA = National Early Inflammatory Arthritis Audit, PtVAS= Patient Global VAS, PhVAS = Physician Global VAS, RAPID3 = Routine Assessment of Patient Index Data 3, RADAI = Rheumatoid Arthritis Disease Activity Index, WPAI = Work Productivity and Activity Impairment, PHQ4ADS = Patient Health Questionnaire 4 item Anxiety and Depression Screener, *Collected but data Not currently published

|  | Part Three | | | | | | | | | | | | | | | | | |
| --- | --- | --- | --- | --- | --- | --- | --- | --- | --- | --- | --- | --- | --- | --- | --- | --- | --- | --- |
| Name and Reference of Registry or Cohort | Treatment History | Clinical Notes | DAS28/JADAS or C/SDAI | RAID | Manifestations/ Features | Steroid Use | Stiffness | T/SJC or A/LJC | Imaging (Radiographs) | MSK-HQ | Pain VAS | EQ-5D | BRAF | Fatigue VAS | (C)HAQ | WSAS | RADS | Treatment Change Reasons |
| Nijmegen Early RA Cohort (Welsing & van Riel, 2004)  (Hiligsmann et al., 2018)* | ✓ | ✓ | ✓ | 🗶 (AIMS) | 🗶 | 🗶 | 🗶 | ✓ | ✓ | 🗶 (AIMS) | ✓ | ✓ | 🗶 (SF-36)* | 🗶 (PtVAS) | ✓ | 🗶 (AIMS & SF-36)* | 🗶 (SF-36)* | 🗶 |
| NoAR (Druce et al., 2015; Symmons & Silman, 2003) | Mainly csDMARD | ✓ | ✓ | 🗶 | 🗶 | ✓ | 🗶 | ✓ | ✓ | 🗶 | ✓ | 🗶 | 🗶 (SF-36) | ✓ | ✓ | 🗶 (SF-36) | 🗶 (SF-36) | 🗶 |
| Nordic JIA Cohort (Nordal et al., 2011) | ✓ | ✓ | ✓ | 🗶 (SF-36) | ✓ | ✓ | 🗶 | ✓ | 🗶 | 🗶 (CHQ) | ✓ | 🗶 (SF-36) | 🗶 (SF-36) | 🗶 (PtVAS) | ✓ | 🗶 (SF-36) | 🗶 (SF-36) | 🗶 |
| NoRDMARD (Olsen et al., 2014) | ✓ | ✓ | ✓ | ✓ | 🗶 | ✓ | 🗶 | ✓ | ✓ | 🗶 | ✓ | ✓ | 🗶 | ✓ | ✓ | 🗶 (WPAI) | 🗶 | 🗶 |
| NPRD (Minden et al., 2002) | Mainly csDMARD | ✓ | 🗶 (PhVAS) | 🗶 | 🗶 | ✓ | 🗶 | ✓ | 🗶 | 🗶 | ✓ | 🗶 | 🗶 | 🗶 (PtVAS) | ✓ | 🗶 | 🗶 | 🗶 |
| ORAR (Kvien & Uhlig, 2004) | ✓ | ✓ | ✓ | 🗶 (AIMS2 & SF-36) | ✓ | 🗶 | Not Clear | ✓ | ✓ | 🗶 (AIMS2 & ASES) | ✓ | 🗶 | 🗶 (SF-36) | ✓ | ✓ | 🗶 (AIMS2 & SF-36) | 🗶  (RAI, AIMS2 & SF-36) | 🗶 |
| PHARMACHILD (Beukelman, Anink, et al., 2017) | Not Clear | Not Clear | ✓ | 🗶 | ✓ | Not clear | Not Clear | ✓ | ✓ | 🗶 | ✓ | 🗶 | 🗶 | 🗶 (PtVAS) | 🗶 | 🗶 | 🗶 | Not clear |

Please Note: T/SJC = Tender/Swollen Joint Count, A/LJC = Active/Limited Joint Count, DAS28 = Disease Activity Score-28 Joint count, JADAS = Juvenile Arthritis Disease Activity Score, C/SDAI = Clinical/Simplified Disease Activity Index, RAID = Rheumatoid Arthritis Impact of Disease, (C)HAQ = (Child) Health Assessment Questionnaire , MSK-HQ = Musculoskeletal Health Questionnaire, VAS = Visual Analogue Scale, EQ-5D = EuroQol 5 Dimensions, BRAF = Bristol Rheumatoid Arthritis Fatigue, WSAS = Work and Social Adjustment Scale, RADS = Rheumatoid Arthritis Distress Scale, NoAR = Norfolk Arthritis Register, NoRDDMARD = Norwegian Antirheumatic Drug Register, NPRD = National Paediatric Rheumatology Database, ORAR = Oslo Rheumatoid Arthritis Register, PHARMACHILD = Pharmacovigilance in JIA patients treated with biologic agents and/or MTX, AIMS (2) = Arthritis Impact Measurement Scales (2), PtVAS = Patient Global VAS, SF-36 = Short Form – 36 Item, CHQ = Child Health Questionnaire, WPAI = Work Productivity and Activity Impairment, PhVAS = Physician Global VAS, RAI = Rheumatology Attitude Index, AIMS2 = Arthritis Impact Measurement Scales 2, SF-36 = Short Form – 36 Item, *Collected but data Not currently published

|  | Part Three | | | | | | | | | | | | | | | | | |
| --- | --- | --- | --- | --- | --- | --- | --- | --- | --- | --- | --- | --- | --- | --- | --- | --- | --- | --- |
| Name and Reference of Registry or Cohort | Treatment History | Clinical Notes | DAS28/JADAS or C/SDAI | RAID | Manifestations/ Features | Steroid Use | Stiffness | T/SJC or A/LJC | Imaging (Radiographs) | MSK-HQ | Pain VAS | EQ-5D | BRAF | Fatigue VAS | (C)HAQ | WSAS | RADS | Treatment Change Reasons |
| QUEST-RA (Sokka et al., 2007) | ✓ | ✓ | ✓ | 🗶 | ✓ | ✓ | ✓ | ✓ | 🗶 | 🗶 | ✓ | 🗶 | 🗶 | ✓ | ✓ | 🗶 | 🗶 | 🗶 |
| RABBIT (Gerhold et al., 2015) | ✓ | ✓ | ✓ | 🗶 (SF-36) | 🗶 | 🗶 | 🗶 | ✓ | 🗶 | 🗶 (SF-36) | ✓ | 🗶 | 🗶 (SF-36) | ✓ | 🗶 (HFSQ | 🗶 (SF-36) | 🗶 (SF-36) | 🗶 |
| RADIUS1 (Gibofsky et al., 2011) | ✓ | ✓ | ✓ | 🗶 | 🗶 | ✓ | 🗶 | ✓ | 🗶 | 🗶 | ✓ | 🗶 | 🗶 | 🗶 (PtVAS) | ✓ | 🗶 | 🗶 | ✓ |
| ReACCh Out (Oen et al., 2009) | ✓ | ✓ | 🗶 | 🗶 (JAQQ) | ✓ | ✓ | 🗶 | ✓ | 🗶 | 🗶 but JAQQ abd QMLQ | ✓ | 🗶 | 🗶 | 🗶 (PtVAS) | ✓ | 🗶 (JAQQ) | 🗶 (JAQQ) | 🗶 |
| REACH (Alves et al., 2011; Kuijper et al., 2014) | ✓ | ✓ | ✓ | 🗶 | 🗶 | 🗶 | 🗶 | ✓ | 🗶 | 🗶 (MHLC & SF-36) | 🗶 (RADAI) | 🗶 | 🗶 (FAS) | ✓ | ✓ | 🗶 (SF-36) | 🗶 (SF-36, CORS & HADS) | 🗶 |
| REUMA.PT (Canhão et al., 2011) | ✓ | ✓ | ✓ | 🗶 | ✓ | 🗶 | 🗶 | ✓ | ✓ | 🗶 (SF-36) | ✓ | 🗶 | 🗶 (SF-36) | 🗶 | ✓ | 🗶 (SF-36) | 🗶 (SF-36) | ✓ |
| Rhumadata© (Choquette et al., 2019) | ✓ | ✓ | ✓ | 🗶 | 🗶 | ✓ | ✓ | ✓ | 🗶 | 🗶 | ✓ | 🗶 | 🗶 | ✓ | ✓ | 🗶 | 🗶 | ✓ |
| ROB-FIN (Nordström et al., 2006) | ✓ | ✓ | ✓ | 🗶 | 🗶 | ✓ | 🗶 | ✓ | 🗶 | 🗶 | ✓ | 🗶 | 🗶 | 🗶 (PtVAS) | ✓ | 🗶 | 🗶 | ✓ |

Please Note: T/SJC = Tender/Swollen Joint Count, A/LJC = Active/Limited Joint Count, DAS28 = Disease Activity Score-28 Joint count, JADAS = Juvenile Arthritis Disease Activity Score, C/SDAI = Clinical/Simplified Disease Activity Index, RAID = Rheumatoid Arthritis Impact of Disease, (C)HAQ = (Child) Health Assessment Questionnaire , MSK-HQ = Musculoskeletal Health Questionnaire, VAS = Visual Analogue Scale, EQ-5D = EuroQol 5 Dimensions, BRAF = Bristol Rheumatoid Arthritis Fatigue, WSAS = Work and Social Adjustment Scale, RADS = Rheumatoid Arthritis Distress Scale, QUEST-RA = Quantitative Patient Questionnaires in Standard Monitoring of Patients with Rheumatoid Arthritis, RABBIT = German register for long-term observation of biologics in RA, RADIUS1 = Rheumatoid Arthritis Disease-Modifying Antirheumatic Drug Intervention and Utilization Study Cohort1, ReACCh Out = Research in Arthritis in Canadian Children emphasizing Outcomes, ROB-FIN = National Register of Biological Treatment in Finland, SF-36 = Short Form – 36 Item, HFSQ = Hannover Functional Status Questionnaire, PtVAS = Patient Global VAS, JAQQ = Juvenile Arthritis Quality of Life Questionnaire, QMLQ = Quality of My Life Questionnaire, MHLC = Multidimensional Health Locus of Control, RADAI = Rheumatoid Arthritis Disease Activity Index, FAS = Fatigue Assessment Scale.

|  | Part Three | | | | | | | | | | | | | | | | | |
| --- | --- | --- | --- | --- | --- | --- | --- | --- | --- | --- | --- | --- | --- | --- | --- | --- | --- | --- |
| Name and Reference of Registry or Cohort | Treatment History | Clinical Notes | DAS28/JADAS or C/SDAI | RAID | Manifestations/ Features | Steroid Use | Stiffness | T/SJC or A/LJC | Imaging (Radiographs) | MSK-HQ | Pain VAS | EQ-5D | BRAF | Fatigue VAS | (C)HAQ | WSAS | RADS | Treatment Change Reasons |
| RRBR (Codreanu et al., 2014) | ✓ | ✓ | ✓ | 🗶 | 🗶 | 🗶 | 🗶 | ✓ | ✓ | 🗶 | 🗶 (PtVAS) | ✓ | 🗶 | 🗶 (PtVAS) | ✓ | 🗶 (Work Productivity) | 🗶 | ✓ |
| SCQM, Switzerland,  (Uitz et al., 2000) | ✓ | ✓ | ✓ | 🗶 | 🗶 | 🗶 | ✓ | ✓ | ✓ | 🗶 | ✓ | 🗶 | 🗶 | 🗶 (PtVAS) | ✓ | 🗶 | 🗶 | 🗶 |
| SRQ (Eriksson et al., 2014)  (Hiligsmann et al., 2018)* | ✓ | ✓ | ✓ | 🗶 | 🗶 | ✓ | 🗶 | ✓ | 🗶 | 🗶 | 🗶 (PtVAS) | ✓ | 🗶 | 🗶 (PtVAS) | ✓ | 🗶 (WAI)* | 🗶 (mFABQ)* | ✓ |
| STURE (Askling et al., 2006) | ✓ | ✓ | ✓ | 🗶 | 🗶 | ✓ | 🗶 | ✓ | 🗶 | 🗶 | ✓ | 🗶 | 🗶 | 🗶 (PtVAS) | ✓ | 🗶 | 🗶 | Not clear |
| Swedish JIA Register (Beukelman, Anink, et al., 2017) | Not Clear | Not Clear | ✓ | No | ✓ | Not clear | Not Clear | ✓ | 🗶 | 🗶 | ✓ | 🗶 | 🗶 | 🗶 (PtVAS) | ✓ | 🗶 | 🗶 | Not clear |
| TURKBIO (Önen et al., 2022) | ✓ | Some | ✓ | 🗶 | 🗶 | ✓ | 🗶 | ✓ | ✓ | 🗶 | ✓ | 🗶 (SF-36) | 🗶 (SF-36) | 🗶 (SF-36) | ✓ | 🗶 (SF-36) | 🗶 (SF-36) | ✓ |
| VARAR (Mikuls et al., 2010) | ✓ | ✓ | ✓ | 🗶 (RAPID3) | ✓ | ✓ | 🗶 | ✓ | 🗶 | 🗶 | ✓ | 🗶 | 🗶 | 🗶 | ✓ | 🗶 | 🗶 | 🗶 |
| WC (Kovalchik et al., 2012) | Mainly csDMARD | ✓ | ✓ | 🗶 (PSH) | ✓ | 🗶 | ✓ | ✓ | ✓ | 🗶 (PSH) | ✓ | 🗶 | 🗶 | ✓ | ✓ | 🗶 (PSH) | 🗶 (PSH) | 🗶 |

Please Note: T/SJC = Tender/Swollen Joint Count, A/LJC = Active/Limited Joint Count, DAS28 = Disease Activity Score-28 Joint count, JADAS = Juvenile Arthritis Disease Activity Score, C/SDAI = Clinical/Simplified Disease Activity Index, RAID = Rheumatoid Arthritis Impact of Disease, (C)HAQ = (Child) Health Assessment Questionnaire , MSK-HQ = Musculoskeletal Health Questionnaire, VAS = Visual Analogue Scale, EQ-5D = EuroQol 5 Dimensions, BRAF = Bristol Rheumatoid Arthritis Fatigue, WSAS = Work and Social Adjustment Scale, RADS = Rheumatoid Arthritis Distress Scale, ROB-FIN = National Register of Biological Treatment in Finland, RRBR = Romanian Registry of Rheumatic Diseases, SCQM = Swiss Clinical Quality Management in Rheumatic Diseases, SRQ = Swedish Rheumatology Quality Registries, STURE = Stockholm TNF follow-Up Registry, TURKBIO = Turkish Biologics Register, VARAR = Veterans Affairs Rheumatoid Arthritis Registry, WC = Western Consortium of Practicing Rheumatologists, PtVAS = Patient Global VAS, WAI = Work Ability Index , mFABQ = modified Fear Avoidance-Belief Questionnaire, SF-36 = Short Form – 36 Item, RAPID3 = Routine Assessment of Patient Index Data 3, PSH = Patient overall satisfaction with health, *Collected but data Not currently publishe
